# Changing patterns in reporting and sharing of review data in systematic reviews with meta-analysis of the effects of interventions: a meta-research study

**DOI:** 10.1101/2022.04.11.22273688

**Authors:** Phi-Yen Nguyen, Raju Kanukula, Joanne E McKenzie, Zainab Alqaidoom, Sue E Brennan, Neal R Haddaway, Daniel G Hamilton, Sathya Karunananthan, Steve McDonald, David Moher, Shinichi Nakagawa, David Nunan, Peter Tugwell, Vivian A Welch, Matthew J Page

## Abstract

**Objectives:** To examine changes in completeness of reporting and frequency of sharing data, analytic code and other review materials in systematic reviews (SRs) over time; and factors associated with these changes.

**Design:** Cross-sectional meta-research study.

**Sample:** A random sample of 300 SRs with meta-analysis of aggregate data on the effects of a health, social, behavioural or educational intervention, which were indexed in PubMed, Science Citation Index, Social Sciences Citation Index, Scopus and Education Collection in November 2020.

**Analysis/Outcomes:** The extent of complete reporting and frequency of sharing review materials in these reviews were compared with 110 SRs indexed in February 2014. Associations between completeness of reporting and various factors (e.g. self-reported use of reporting guidelines, journal’s data sharing policies) were examined by calculating risk ratios (RR) and 95% confidence intervals (CI).

**Results:** Several items were reported sub-optimally among 300 SRs from 2020, such as a registration record for the review (38%), a full search strategy for at least one database (71%), methods used to assess risk of bias (62%), methods used to prepare data for meta-analysis (34%), and funding source for the review (72%). Only a few items not already reported at a high frequency in 2014 were reported more frequently in 2020. There was no evidence that reviews using a reporting guideline were more completely reported than reviews not using a guideline. Reviews published in 2020 in journals that mandated either data sharing or inclusion of Data Availability Statements were more likely to share their review materials (e.g. data, code files) (18% vs 2%).

**Conclusion:** Incomplete reporting of several recommended items for systematic reviews persists, even in reviews that claim to have followed a reporting guideline. Data sharing policies of journals potentially encourage sharing of review materials.

SUMMARY BOX

What is already known on this topic

- Complete reporting of methods and results, as well as sharing data and analytic code, enhances transparency and reproducibility of systematic reviews. The extent of complete reporting and sharing of data or analytic code among systematic reviews needs to be comprehensively assessed.
- Use of reporting guidelines, which are designed to improve reporting in systematic reviews, is increasing. It is unclear whether this increase has had an impact on reporting of methods and results in systematic reviews.
- More journals are adopting open data policies which aim to promote data sharing. The impact of these policies on sharing data and analytic code in systematic reviews is also unclear.

What this study adds

- Incomplete reporting of several recommended items in systematic reviews persists. Frequency of sharing review data and analytic code is currently low (7%).
- An increase in self-reported use of a reporting guideline was observed between 2014-2020; however, there was no evidence that reviews using a reporting guideline were more completely reported than reviews not using a guideline.
- Reviews published in 2020 in journals that mandated either data sharing or inclusion of Data Availability Statements were more likely to share their review materials (e.g. data, code files).

## INTRODUCTION

Systematic reviews underpin many government policies and professional society guideline recommendations [1]. To ensure systematic reviews are valuable to decision makers, authors should completely report the methods and results of their review. Complete reporting allows users to judge whether the chosen methods may have biased the review findings. Incomplete reporting of the methods prevents such an assessment and can preclude attempts to replicate the findings. Several meta-research studies have evaluated the completeness of methods and results reporting in systematic reviews and meta-analyses. Many of these were narrow in scope, focusing only on reviews of specific health topics [2–6] or reviews published in selected journals [7,8]. In other studies, the examined sample of reviews were relatively more diverse, but were published almost a decade ago [9,10], or were evaluated against a small set of reporting items [1], meaning that comprehensive data on the current state of systematic review reporting is lacking.

To address incomplete reporting of methods and results in systematic reviews, several reporting guidelines have been developed, with the Preferred Reporting Items for Systematic reviews and Meta-Analyses (PRISMA) statement [11] among the more widely used [12]. Reporting guidelines provide a structure for reporting a systematic review, along with recommendations of items to report [13]. Originally released in 2009, PRISMA was recently updated (to PRISMA 2020) to reflect advances in systematic review methodology [14]. The few studies examining the impact of PRISMA suggest that for some items (e.g. inclusion of a flow diagram) there was improvement after the introduction of the 2009 PRISMA statement, but that others (e.g. mention of a review protocol) remained infrequently reported [15]. Such evaluations are limited to reviews in particular health areas published prior to 2015 and so it is unclear whether reporting guidelines have had an influence on more a recent, general sample of systematic reviews.

In addition to transparent reporting, advocates for research transparency [16,17] also recommend authors share systematic review data files and analytic code used to generate meta-analyses [18]. While all data for a meta-analysis is typically summarised in a tables or forest plots, sharing an editable file containing extracted data (e.g., CSV, RevMan) reduces the time and risk of errors associated with manual extraction of such data. This then facilitates the review’s reuse in future updates and replications, or its inclusion in overviews of reviews, clinical practice guidelines, educational materials and meta-research studies [18,19]. Sharing of review data files is relatively easier than sharing individual participant data from primary studies and signals that review authors are committed to practices they like to see performed by authors of primary studies, who are often requested to share their data. Infrequent sharing of data in systematic reviews in health research has been observed, but these findings may not generalise to all health topics [4] or across journals [7]. Moreover, the types of data shared (e.g. unprocessed data extracted from reports, data included in meta-analyses) has not been examined, nor has the impact of journal data sharing policies on sharing rates in systematic reviews.

Without a current, comprehensive, evaluation of the completeness of reporting of systematic reviews, we lack data on which items are infrequently reported and require most attention from authors, peer reviewers, editors and educators. Furthermore, without data on the frequency and type of materials review authors currently share, we lack insight into how receptive review authors are to calls to share data underlying research projects. To address these research gaps, we aimed to:

a. evaluate the completeness of reporting in systematic reviews in a cross-section of systematic reviews with meta-analysis published in 2020;
b. evaluate the frequency of sharing review data, analytic code and other materials in the same cohort of reviews;
c. compare reporting in these reviews with a sample of reviews published in 2014;
d. investigate the impact of reporting guidelines on the completeness of reporting of reviews published in 2020; and investigate the impact of data sharing policies of journals on the frequency of review data sharing in reviews published in 2020.
e. We chose 2014 as the year to compare reviews from 2020 against because we had access to the raw data on reporting completeness in a sample of reviews from 2014 [10] that met the same eligibility criteria and were evaluated using similar methods as the reviews sampled from 2020.

## METHODS

This study was conducted as one of a suite of studies in the REPRISE (REProducibility and Replicability In Syntheses of Evidence) project. The REPRISE project is investigating various aspects relating to the transparency, reproducibility and replicability of systematic reviews with meta-analysis of the effects of health, social, behavioural and educational interventions. Methods for all studies were pre-specified in the same protocol [20]. Deviations from the protocol for the current study are outlined in Supplemental data.

### Identification and selection of articles

We included a random sample of systematic reviews with meta-analysis of the effects of a health, social, behavioural or educational intervention (that is, any intervention designed to improve health [defined as “a state of complete physical, mental and social well-being and not merely the absence of disease or infirmity” [21], promote social welfare and justice, change behaviour or improve educational outcomes; see Supplemental data for full eligibility criteria). To be considered a “systematic review”, authors needed to have, at a minimum, clearly stated their review objective(s) or question(s); reported the source(s) (e.g. bibliographic databases) used to identify studies meeting the eligibility criteria; and reported conducting an assessment of the validity of the findings of the included studies, for example via an assessment of risk of bias or methodological quality. We did not exclude systematic reviews providing limited detail about the methods used. We only included systematic reviews that presented results for at least one pairwise meta-analysis of aggregate data. Systematic reviews with network meta-analyses were eligible if they included at least one direct (i.e. pairwise) comparison that fulfilled the above-mentioned criteria. Systematic reviews with only meta-analyses of individual participant data (IPD) were excluded because all eligible systematic reviews in this study will be subjected to a reproducibility check in another component of the REPRISE project [20], and we lack the resources to reproduce IPD meta-analyses. Furthermore, only reviews written in English were included.

Using search strategies created by an information specialist (SM), we systematically searched PubMed, Science Citation Index (SCI) and Social Sciences Citation Index (SSCI) via Web of Science, Scopus via Elsevier and Education Collection via ProQuest for systematic reviews indexed from November 2nd to December 2nd, 2020. All searches were conducted on December 3rd, 2020. An example of the search strategy for PubMed was: (meta-analysis[PT] OR meta-analysis[TI] OR systematic[sb]) AND 2020/11/02:2020/12/02[EDAT]. Search strategies for all databases are available in Supplemental data.

We used Endnote v9.3.3 for automatic deduplication of records, then randomly sorted unique records in Microsoft Excel using the RAND() function, and imported the first 2,000 records yielded from the search into Covidence [22] for screening. Two authors (MJP and either PN or RK) independently screened the titles and abstracts of the 2,000 records against the eligibility criteria. We retrieved the full text of all records deemed potentially eligible, and two authors (PN and either MJP or RK) independently evaluated them in random order against the eligibility criteria until we reached our target sample size of 300 systematic reviews. Any disagreement at each stage of screening was resolved via discussion or adjudication by the senior reviewer (MJP). As this was primarily a descriptive study, our aim was to examine reporting across a range of practices. We selected our sample size of 300 systematic reviews as a balance of feasibility and precision. This sample size allowed us to restrict the width of a 95% two-sided Wald type confidence interval around the estimated percentage of reviews reporting a particular practice to a *maximum* of 6%, assuming a prevalence of 50%. For a prevalence of less (or greater) than 50%, the absolute width will be smaller. This maximum confidence interval width was small enough such that our interpretation of the confidence interval limits would generally be consistent.

## Data collection

Two authors (PN and either MJP, RK or ZA) collected data independently and in duplicate from all of the 300 systematic reviews using a standardised form created in REDCap v10.6.12, hosted at Monash University [23]. Any disagreement in the data collected was resolved via discussion or adjudication by the senior reviewer (MJP). Prior to data collection, a pilot test of the data collection form was performed on a random sample of 10 systematic reviews and the form was adjusted as necessary. The full data collection form (Supplemental data) includes a subset of items used in previous evaluations of completeness of reporting [9,10] along with additional items to capture some issues not previously examined. The wording of items in the data collection form was matched to previous evaluations [9,10] to facilitate comparison.

The form consisted of three sections (Table 1). The first section captured general characteristics of the review, which were all extracted manually, except for the country of the corresponding author, which was extracted using R code adapted from the *easyPubMed* package v2.13 [24,25]. The interventions were classified as health, behavioural, social or educational interventions (see definitions in Supplemental data). The second section consisted of items describing reporting characteristics of the review and the index meta-analysis (defined as the first meta-analysis mentioned in the Abstract/Results sections), and data-sharing characteristics of the review. All of the reporting items evaluated are recommended in the 2009 PRISMA statement (either in the main checklist or explanation and elaboration document [26]), except for the items on whether search strategies for *all* bibliographic databases and non-database sources were reported. To facilitate our analysis of the impact of reporting guidelines, we also recorded whether the authors self-reported using a reporting guideline, defined as any document specifying essential items to report in a systematic review (e.g. PRISMA, MECIR or MECCIR standards, etc.)

**Table 1.**
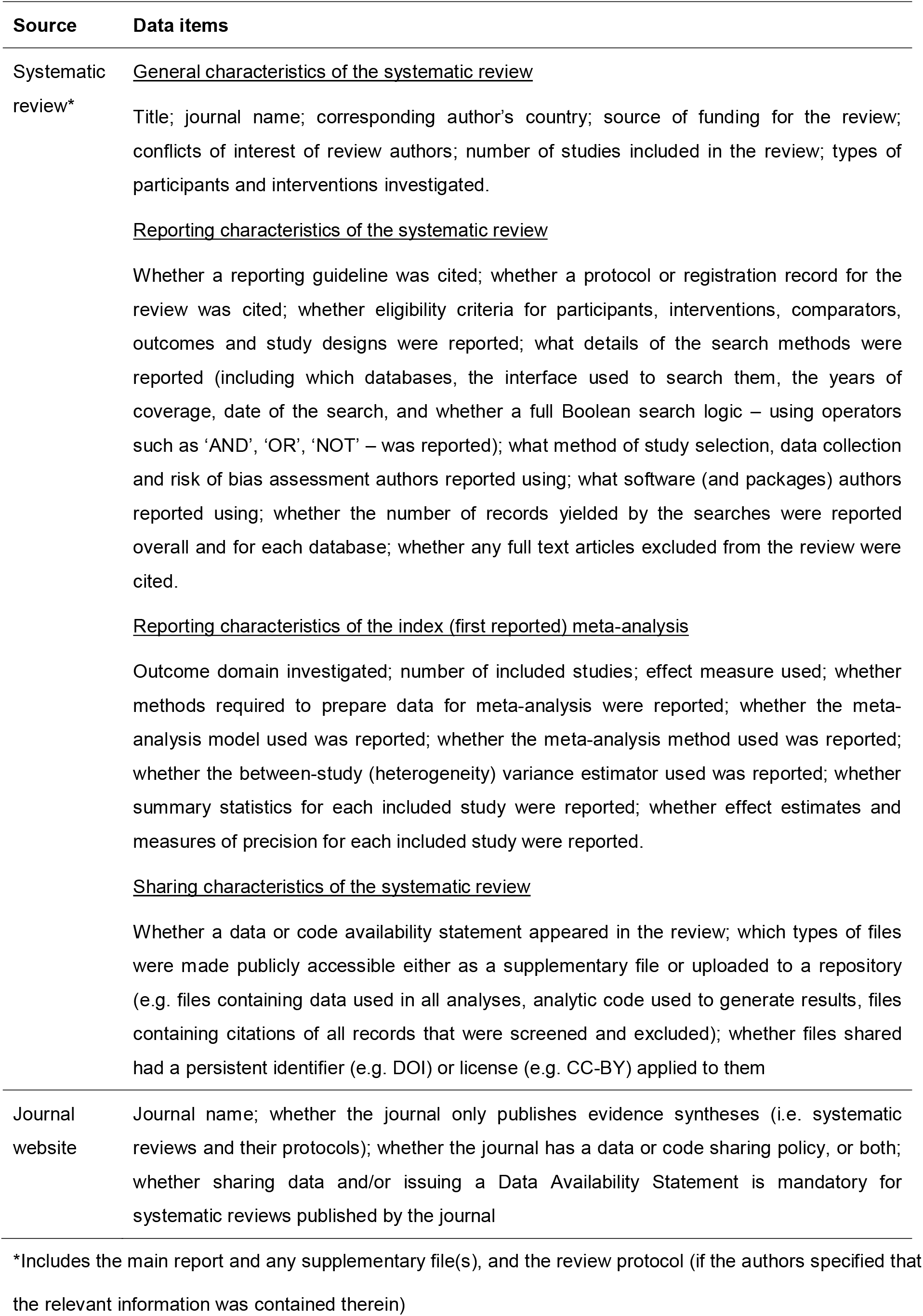
Items for data collection and data sources (see S4 Appendix for further details)

The final section captured the data sharing policy of the journal where the article was published. A data sharing policy refers to the journal’s requirements and expectations regarding public sharing of data and code used in the review. Web archives (https://web.archive.org/) were used to retrieve the version of the policy published prior to 1 November 2020.

We collected data from the main report of the systematic review, any supplementary file provided on the journal server or any cited repository, the review protocol if the authors specified that the relevant information was contained therein, as well as journal websites (Table 1). In the event of discrepancies between the protocol and the main report, we gave preference to data from the main report.

### Secondary use of data collected on systematic reviews from 2014

We obtained the dataset previously collated by Page et al. [10], which included data on completeness of reporting and sharing of review data in a random sample of 110 systematic reviews of health interventions indexed in MEDLINE in February 2014. The reviews included in the 2014 dataset were drawn from a random sample of 300 systematic reviews of health research that addressed questions of intervention efficacy, diagnostic test accuracy, epidemiology or prognosis, 110 of which evaluated the effect of health interventions and met the same eligibility criteria that the 2020 reviews had to meet (apart from year of publication). We extracted from the 2014 dataset individual review data for all reporting and sharing items that were worded the same or similarly as the items collected in the 2020 sample. Where necessary, we recoded data in the 2014 sample to ensure harmonisation with the 2020 sample. We did not collect any additional data on the systematic reviews (or the journals they were published in) in the 2014 sample. Given the systematic reviews in 2014 were identified via MEDLINE only, whereas the systematic reviews in 2020 were identified via five databases (PubMed, SCI, SSCI, Scopus and Education Collection), we determined how many of the included reviews from 2020 happened to also be indexed in MEDLINE, to ensure the comparison between years was appropriate.

## Data analysis

We summarised general and reporting characteristics of the included systematic reviews using descriptive statistics (e.g. frequency and percentage for categorical items, median and interquartile range for continuous items). We calculated risk ratios to quantify differences in the percentage of reviews meeting indicators of ‘completeness of reporting’ and ‘sharing of review materials’ between the following groups:

a. reviews published in 2020 in an evidence synthesis journal (defined as a journal which has a strong or exclusive focus on systematic reviews and their protocols, as identified from the journal website’s Aims and Scope sections) *versus* published elsewhere;
b. reviews of health interventions published in 2020 *versus* reviews of health interventions published in 2014
c. reviews published in 2020 reporting use of a reporting guideline (e.g. PRISMA) *versus* not reporting use;
d. reviews published in 2020 in journals with *versus* without a data-sharing policy;
e. reviews published in 2020 in journals with *versus* without a policy that mandates either data sharing or declaration of data availability, irrespective of whether the policy applies universally to all studies or specifically to systematic reviews.

Risk ratios (RR) and Wald-type normal 95% confidence intervals (CI) were calculated using the *epitool* package v0.5-10.1 (R v4.0.3) [27]. When the numerators were small (<5) in either group, or the outcome was very rare (<5%) in either group, we instead used penalised likelihood logistic regression (implemented via the *logistf* package v1.24 in R [28]. Penalised likelihood logistic regression has been shown to improve estimation of the odds ratio and its confidence interval for rare events or unbalanced samples [29,30]. The odds ratios from these models can be interpreted as risk ratios when the events are rare in both groups [31]. The RRs and their 95% CIs were displayed using forest plots (implemented via the *forestplot* package v1.10.1 in R) [32]. Rather than relying on statistical significance when interpreting RR associations (i.e. claiming that an association exists when the 95% confidence interval did not include the null), we defined an equivalence range for all comparisons as [0.9 to 1.1] – any RR less than 0.9 or more than 1.1 (i.e. a 10% difference in rate of reporting in either direction) was deemed as an important difference. Since no previous study has identified a meaningful threshold for important changes in reporting in systematic reviews, this equivalence range was determined based on consensus between investigators. Assuming an item was reported by 50% of reviews in 2014, a RR of 1.1 reflects that the item was reported by 55% of reviews in 2020 (a difference of 5 percentage points). If the reporting rate in 2014 is higher than 50% (e.g. 80%), the threshold to be considered an important difference will be higher (i.e. 8 percentage points).

We conducted two post-hoc sensitivity analyses, the first by excluding Cochrane reviews because they were subjected to strict editorial processes to ensure adherence to methodological conduct and reporting standards, and the second by excluding reviews on COVID-19 due to concerns about short publication turnarounds, which could have an impact on reporting quality [33].

### Patient and public involvement

We did not directly involve patients or members of the public when we designed our study, interpreted the results or wrote the manuscript, because our focus was to identify problems in how researchers report their work in scientific journals with a predominantly scientific readership. However, the idea for our study arose from our concerns as people who interact with the healthcare system that incomplete reporting can lead to undue trust being placed in the findings of flawed systematic reviews, potentially leading to ineffective or harmful treatments being delivered. We asked a member of the public to read our manuscript after submission to ensure it was understandable.

## RESULTS

### Results of the search

Our search retrieved 8,208 records (Fig. 1). Out of the first 2,000 randomly sorted titles and abstracts that were screened, we considered 603 as potentially eligible and retrieved the full text for screening. We only needed to screen the first 436 randomly sorted full text reports to reach our target sample size of 300. Citations of all records identified, screened, excluded and included are available on the Open Science Framework (DOI: <u>10.17605/OSF.IO/JSP9T</u>).

**Fig. 1.**
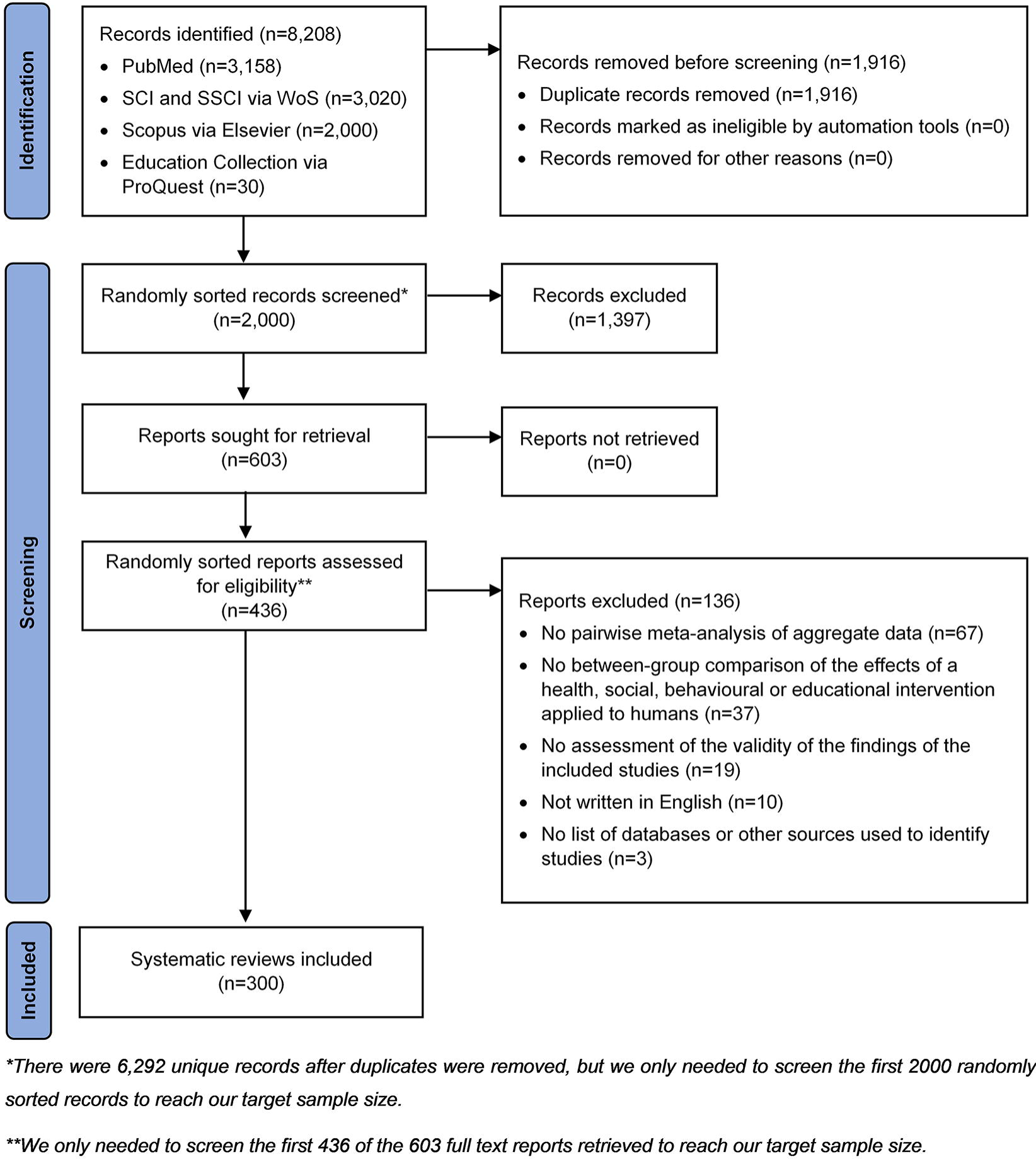
PRISMA 2020 flow diagram of identification, screening and inclusion of systematic reviews

### General characteristics of systematic reviews

Among the 2020 sample (n=300), half of the systematic reviews (n=151, 50%) had a corresponding author based in one of three countries – China (32%), the United States (10%) and United Kingdom (8%) (Table 2). The reviews included a median of 12 studies (IQR 8-21), with index meta-analyses including a median of 6 studies (IQR 4-10). Most reviews (n=215, 72%) included a financial disclosure statement, of which 97 (32%) declared no funding. Most review authors (n=251, 84%) declared having no conflict of interest. Common softwares used for meta-analysis were Review Manager (n=189, 63%), Stata (n=73, 24%) and R (n=33, 11%).

**Table 2.** Descriptive characteristics of systematic reviews indexed in 2020

| Item | Median (IQR) or frequency (%) |
| --- | --- |
| Number of databases searched | 4 (3-5) |
| Number of studies included in review | 12 (8-21) |
| Number of studies in index meta-analysis | 6 (4-10) |
| Country of corresponding author |  |
| China | 96/300 (32) |
| USA | 31/300 (10) |
| UK | 24/300 (8) |
| Other | 149/300 (50) |
| Source of funding |  |
| Non-profit | 112/300 (37) |
| For-profit | 3/300 (1) |
| Both for-profit and non-profit | 3/300 (1) |
| No funding | 97/300 (32) |
| Not reported | 85/300 (28) |
| Conflict of interest |  |
| Present | 30/300 (10) |
| Not present | 251/300 (84) |
| Not declared | 19/300 (6) |
| ICD-11 category investigated |  |
| Endocrine, nutritional or metabolic diseases | 36/300 (12) |
| Diseases of the digestive system | 36/300 (12) |
| Diseases of the musculoskeletal system or connective tissue | 35/300 (12) |
| Diseases of the circulatory system | 30/300 (10) |
| Other | 198/300 (66) |
| Type(s) of intervention |  |
| Pharmacological | 102/300 (34) |
| Non-pharmacological | 189/300 (63) |
| Both | 9/300 (3) |
| Area(s) of intervention |  |
| Health | 294/300 (98) |
| Behavioural | 28/300 (9) |
| Educational | 4/300 (1) |
| Social | 5/300 (2) |
| Citing a reporting guideline | 245/300 (82) |
| **Reporting of the review process** |  |
| Protocol/registration record cited |  |
| Both protocol and registration record cited | 1/300 (<0.5) |
| Only a protocol cited | 13/300 (4) |
| Only a registration record cited | 112/300 (37) |
| Neither | 174/300 (58) |
| Eligibility criteria stated |  |
| Participants | 275/300 (92) |
| Interventions/Exposures | 296/300 (99) |
| Comparators | 267/300 (89) |
| Outcomes | 298/300 (99) |
| Study design | 278/300 (93) |
| Type(s) of eligible study design |  |
| Randomised studies | 168/278 (60) |
| Non-randomised studies | 20/278 (7) |
| Both | 90/278 (32) |
| Search interface reported (e.g. Ovid for MEDLINE) |  |
| For all databases | 76/300 (25) |
| For some databases | 36/300 (12) |
| Not reported | 188/300 (63) |
| Dates of coverage of databases reported |  |
| Both exact start and end dates | 136/300 (45) |
| Both start and end dates but not as exact dates (e.g. "from inception to May 2020") | 105/300 (35) |
| Only start or end date | 50/300 (17) |
| Not reported | 9/300 (3) |
| Exact last date of search reported<sup>#</sup> | 72/300 (24) |
| Search strategy for databases reported |  |
| Boolean logic for all databases | 81/300 (27) |
| Boolean logic for some databases | 133/300 (44) |
| List of MeSH & free text terms only | 12/300 (4) |
| List of MeSH terms only | 8/300 (3) |
| List of free text terms only | 59/300 (20) |
| Not reported | 7/300 (2) |
| Trials register searched | 64/300 (21) |
| Other electronic sources searched | 102/300 (34) |
| Search strategy for trial register/other sources reported | 24/140 (17) |
| Method of screening |  |
| All studies screened by at least 2 authors | 196/300 (65) |
| At least 2 authors were involved in either titles/abstracts or full-text screening, but unclear for the other step | 22/300 (7) |
| Different methods were applied for titles/abstracts and full-text screening | 7/300 (2) |
| All studies screened by 1 author and a subset by another author | 4/300 (1) |
| All studies screened by 1 author with the use of an automation tool | 0/300 (0) |
| All studies screened by 1 author only | 4/300 (1) |
| Not reported | 67/300 (22) |
| Method of data collection |  |
| All data collected by 2 authors | 208/300 (69) |
| All data collected by 1 author with verification by another | 16/300 (5) |
| All data collected by 1 author only | 4/300 (1) |
| Other arrangements | 1/300 (<0.5) |
| Not reported | 71/300 (24) |
| Method of ROB assessment |  |
| All studies assessed by 2 authors | 173/300 (58) |
| All studies assessed by 1 author with verification by another | 7/300 (2) |
| All studies assessed by 1 author only | 5/300 (2) |
| Not reported | 115/300 (38) |
| ROB assessment reported for each study | 231/300 (77) |
| Total records retrieved reported | 300/300 (100) |
| Records retrieved per database reported | 126/300 (42) |
| Software(s) used for meta-analysis |  |
| Review Manager | 189/300 (63) |
| Stata | 73/300 (24) |
| R | 33/300 (11) |
| Comprehensive Meta-Analysis | 27/300 (9) |
| Other | 13/300 (4) |
| SPSS | 4/300 (1) |
| SAS | 1/300 (<0.5) |
| Not reported | 3/300 (1) |
| Software details reported |  |
| Both analysis package and software version reported | 28/297 (9) |
| Only analysis package reported | 8/297 (3) |
| Only software version reported | 242/297 (81) |
| Neither | 19/297 (6) |
| At least 1 excluded article cited | 65/300 (22) |
| **Index meta-analysis** |  |
| Measure of effect used |  |
| Risk ratio | 72/300 (24) |
| Odds ratio | 71/300 (24) |
| Hazard ratio | 13/300 (4) |
| Risk difference | 3/300 (1) |
| Mean difference | 76/300 (25) |
| Standardised mean difference | 63/300 (21) |
| Other | 2/300 (1) |
| Method of data preparation reported | 101/300 (34) |
| Meta-analysis model reported (e.g. fixed-effects, random-effects) | 294/300 (98) |
| Meta-analysis method reported (e.g. Mantel-Haenszel, inverse variance) | 218/300 (73) |
| Heterogeneity variance estimator reported (e.g. DerSimonian-Laird) | 50/235 (21) |
| Summary statistics reported for each study | 215/300 (72) |
| Effect estimate and measure of precision reported for each study | 288/300 (96) |
| **Sharing of data and materials used in analyses** |  |
| Data/code availability statement present | 93/300 (31) |
| Types of data shared |  |
| Unprocessed extracted data | 9/300 (3) |
| Data conversions performed | 1/300 (<0.5) |
| Data used in analyses | 12/300 (4) |
| Analytic code | 2/300 (1) |
| Citations of all screened studies | 2/300 (1) |
| Metadata of shared files | 1/300 (<0.5) |
| Any of the above | 20/300 (7) |
| Method(s) of sharing |  |
| Supplementary files | 20/20 (100) |
| Open-access repository | 1/20 (5) |
| Institutional repository | 1/20 (5) |
| DOI cited for shared data | 2/2 (100) |
| License stated for shared data | 2/2 (100) |
| **Journal that publishes the review** |  |
| Specialised in evidence syntheses (e.g. Cochrane Database of Systematic Reviews) | 14/300 (5) |
| Data policy stated in guideline for authors | 199/300 (66) |
| Sharing of data and/or analytic code is encouraged, but it is not mandatory for publication in the journal, and a Data Availability Statement is not required. | 112/199 (56) |
| Sharing data and/or analytic code is not mandatory for publication of systematic reviews, but a Data Availability Statement, which contains links to shared data or reasons for not sharing data, must be provided. | 51/199 (26) |
| Sharing data and/or analytic code is a condition of publication of systematic reviews by the journal. | 36/199 (18) |

\*We additionally recorded whether the author confirmed the date of the last search, which, in practice, may or may not be the same as from the end date of the search range.

The included reviews covered a wide range of topics. The intervention was classified as a health intervention in nearly all reviews (n=294, 98%), and as a behavioural, social or educational intervention in 37 (12%) of reviews (some reviews examined both types of interventions). Almost two-thirds of the reviews (n=198, 66%) examined the effects of non-pharmacological interventions. Out of 24 ICD-11 categories of diseases and conditions, our sample of reviews captured 23 categories. The top four categories (endocrine, nutritional or metabolic diseases, diseases of the digestive system, the musculoskeletal system, and the circulatory system) accounted for 46% of all systematic reviews.

The included systematic reviews were published across 223 journals. Five journals (accounting for 5% of all systematic reviews) specialised in evidence synthesis; 140 journals (accounting for 66% of all systematic reviews) outline a data-sharing policy in the instruction page for authors (Supplemental data).

The general characteristics of the 2014 sample (n=110) have been described elsewhere [10]. In brief, the 2014 sample was similar to the 2020 sample in many aspects, such as the sample size of each review (median=13 studies, IQR 7-23), size of the index meta-analysis (median=6 studies, IQR 3-11) and the prevalence of non-pharmacological reviews (n=55, 50%). Like the 2020 sample, the reviews in 2014 were published in a wide range of journals (n=63), addressed several clinical topics (19 ICD-10 categories) and predominantly had corresponding authors from China, the UK and Canada (n=55 combined, 50%).

### Completeness of reporting of reviews in systematic reviews from 2020

Of the items we examined, the most frequently reported included the total number of records yielded from searches (100%), a declaration of review authors’ conflicts of interest (94%), each of the PICOS (Participants, Interventions, Comparators, Outcomes and Study designs) components of the eligibility criteria (89-99%), the meta-analysis model (e.g. fixed-effect) used (98%) and the effect estimates, together with the measures of precision, for each study included in the index meta-analysis (96%) (Table 2). On the other hand, several items were reported in between 50% to 80% of reviews. These included the funding source for the review (72%), start and end dates of coverage of databases searched (80%), a full Boolean search logic for some or all databases (71%), methods used to screen studies (78%), methods used to collect data (76%), methods used to assess risk of bias (62%), the meta-analysis method (e.g. Mantel-Haenszel, inverse variance) used (73%), and summary statistics for each study included in the index meta-analysis (72%).

Several items were reported in fewer than 50% of reviews. These included a registration record (38%) or protocol (4%) for the review, the interfaces used to search databases (e.g. Ovid, EBSCOhost) (37%), a search strategy for sources that are not bibliographic databases (17%), number of records retrieved for each database (42%), citation for at least one excluded article (22%), methods of data preparation (e.g. data conversion, calculation of missing statistics) (34%) and the heterogeneity variance estimator used for the index meta-analysis (21%).

### Sharing of data, analytic code and other review materials in systematic reviews from 2020

In our 2020 sample, 20 systematic reviews (7%) made data files or analytic code underlying the meta-analysis publicly available, which included two reviews (1%) that shared analytic code. All of these reviews shared these data via supplementary files; two reviews additionally hosted data and analytic code in a public repository. The most commonly shared materials were data files used in analyses, such as RevMan (.rm5) files (n=12/20).

### Changing patterns of reporting between 2014-2020

Of the 300 systematic reviews from 2020, 294 were systematic reviews of health interventions, which we compared with 110 reviews of health interventions from 2014. We determined that 87% of the 294 reviews from 2020 were indexed in MEDLINE; given this high percentage, we consider the comparison with systematic reviews indexed in MEDLINE in 2014 to be appropriate. Compared to the 2014 reviews, systematic reviews indexed in 2020 more frequently cited a reporting guideline (82% vs 29%), were more likely to report a full search strategy for at least one database (72% vs 55%), the total number of records retrieved (100% vs 83%) and data preparation methods (34% vs 15%); 95% CIs for all risk ratios exceeded the upper limit of the equivalence range (Fig 2). For six reporting items, frequencies in both years were similarly high (>90%), leaving little room for improvement. For six other reporting items, frequency of reporting in both years was less than 80% and the estimated differences between years were uncertain as the 95% CIs included the equivalence range (Fig. 2). In a sensitivity analysis excluding Cochrane reviews from both samples (Supplemental data), some existing differences became more pronounced, or 95% confidence intervals narrowed.

**Fig. 2.**
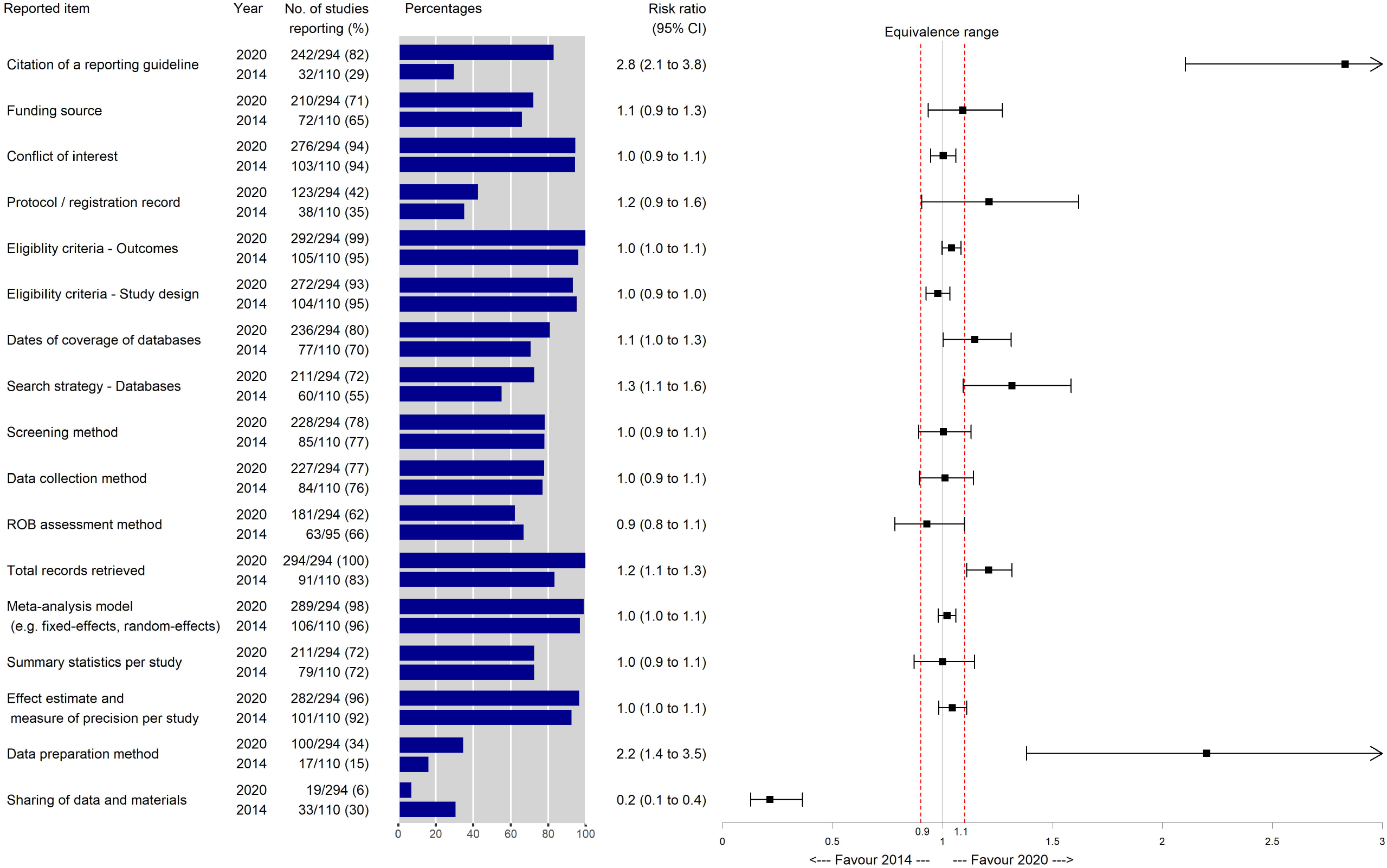
Frequency of reporting items between systematic reviews indexed in 2014 and 2020

### Impact of reporting guidelines, journal type and data sharing policies on reporting in systematic reviews from 2020

Of the 300 reviews, 245 (82%) reported using a reporting guideline. There was no evidence that such reviews were more completely reported than reviews not using a guideline, as for all reporting items, 95% CIs for the risk ratios crossed the equivalence range (Fig. 3A-B). However, of the 27 reporting items compared, nine were reported at a high frequency (>90%) in both groups, leaving little opportunity for a difference. We conducted a sensitivity analysis by excluding systematic reviews on COVID-19 (n=6) from both groups, but no notable changes were observed (Supplemental data).

**Fig. 3A.**
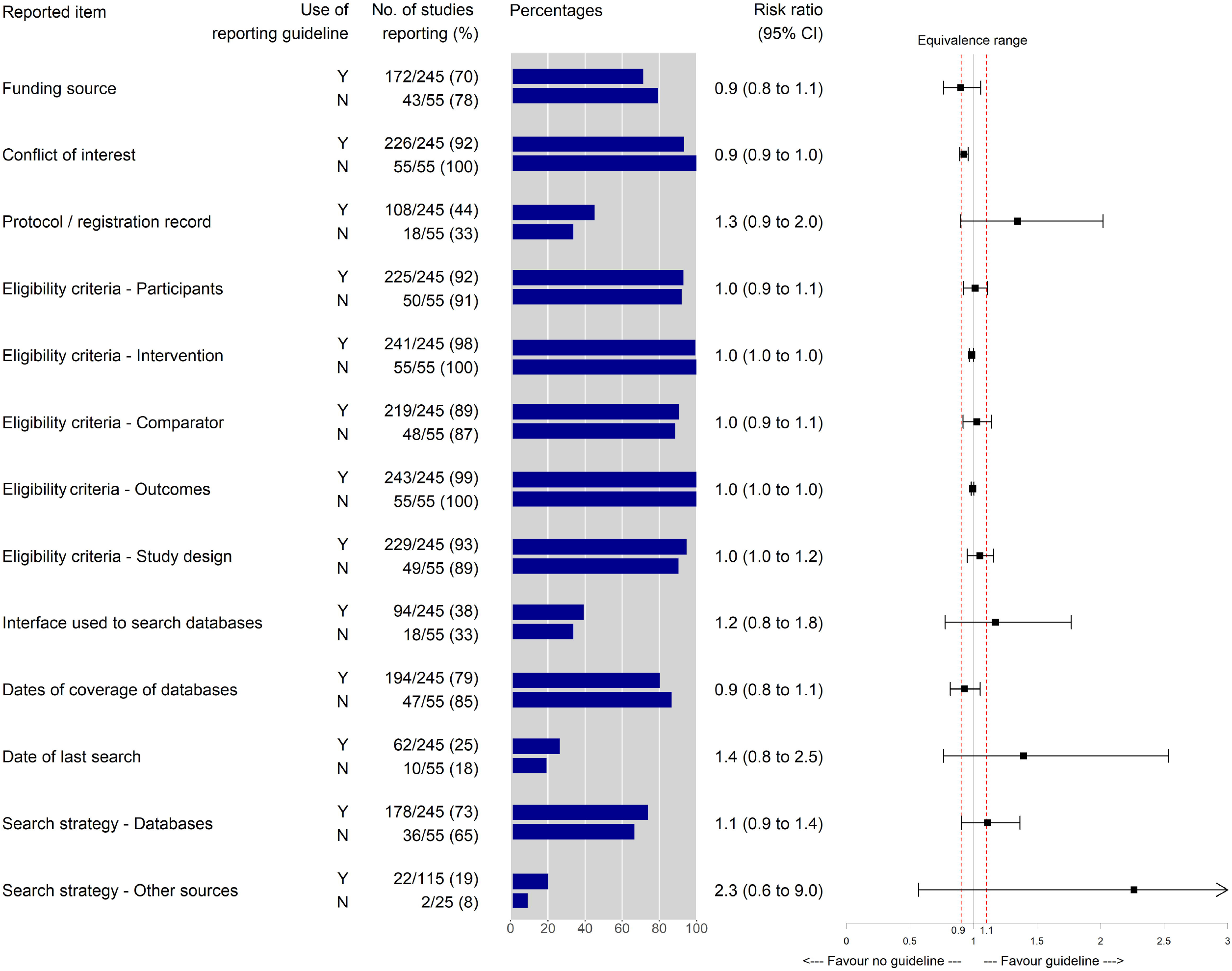
Relationship between citation of a reporting guideline and reported items

**Fig. 3B.**
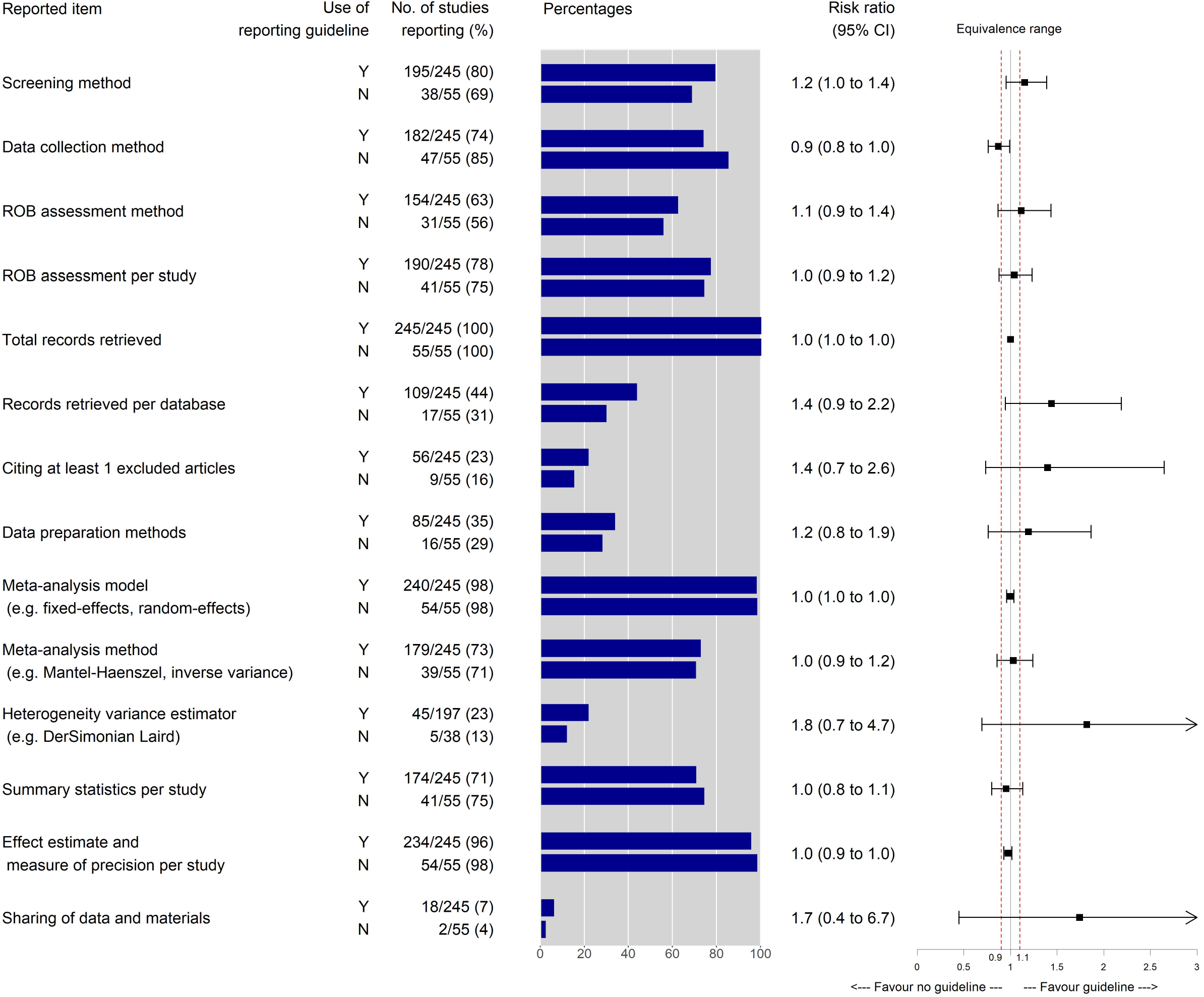
Relationship between citation of a reporting guideline and reported items (ctn’d)

Only 14 systematic reviews from 2020 were published in specialist evidence synthesis journals, including eight Cochrane reviews. Such reviews were reported more completely than reviews published elsewhere, with 95% CIs for risk ratios exceeding the upper limit of the equivalence range for 13 of 28 reporting items compared (Fig. 4A-B). Such items included those that have received limited attention in previous meta-research studies, such as the interface used to search bibliographic databases (79% vs 35%), a search strategy for non-database sources (78% vs 13%), citation for at least one excluded study (64% vs 20%) and availability of data and materials (57% vs 4%).

**Fig. 4A.**
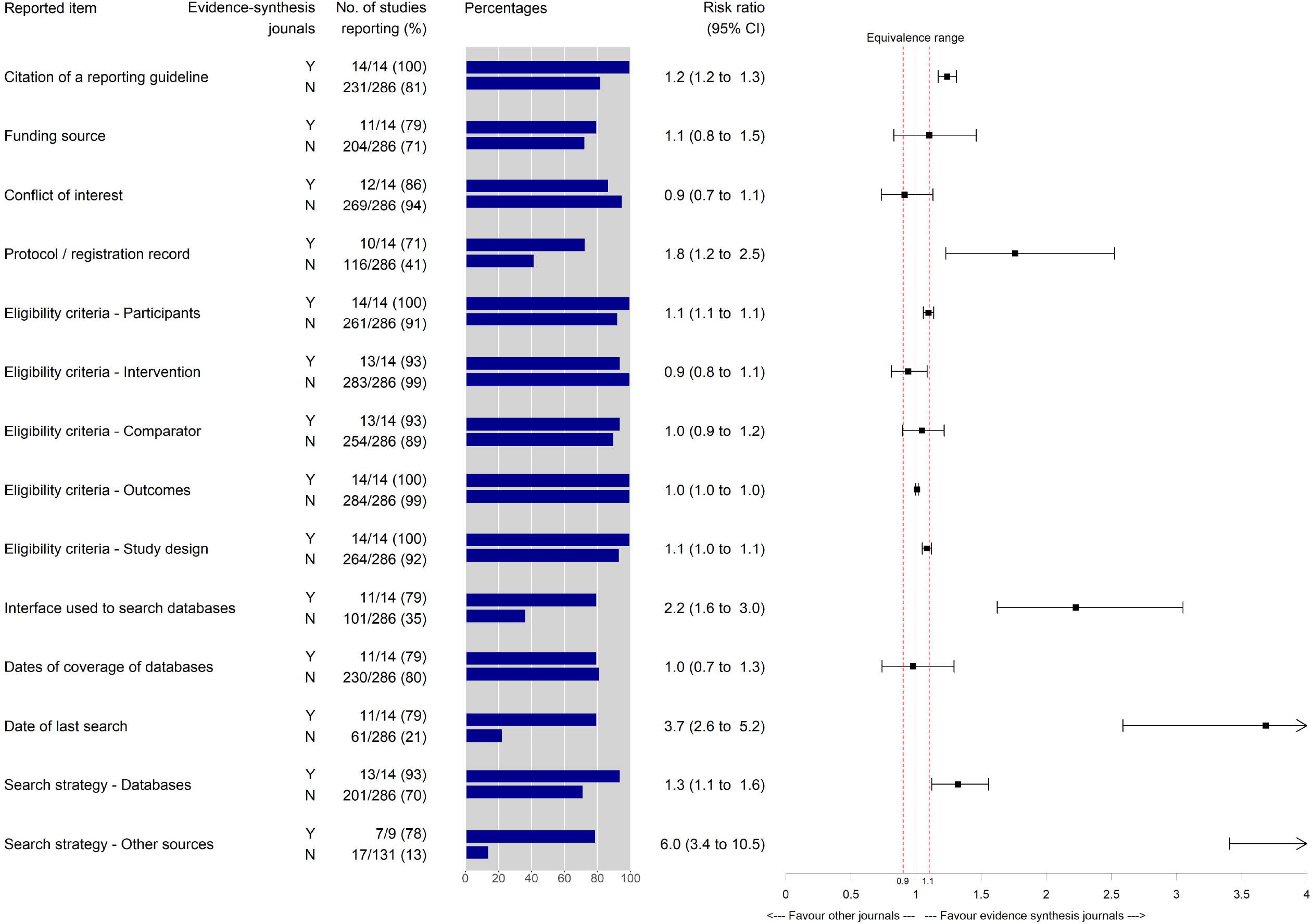
Relationship between journal type and reported items

**Fig. 4B.**
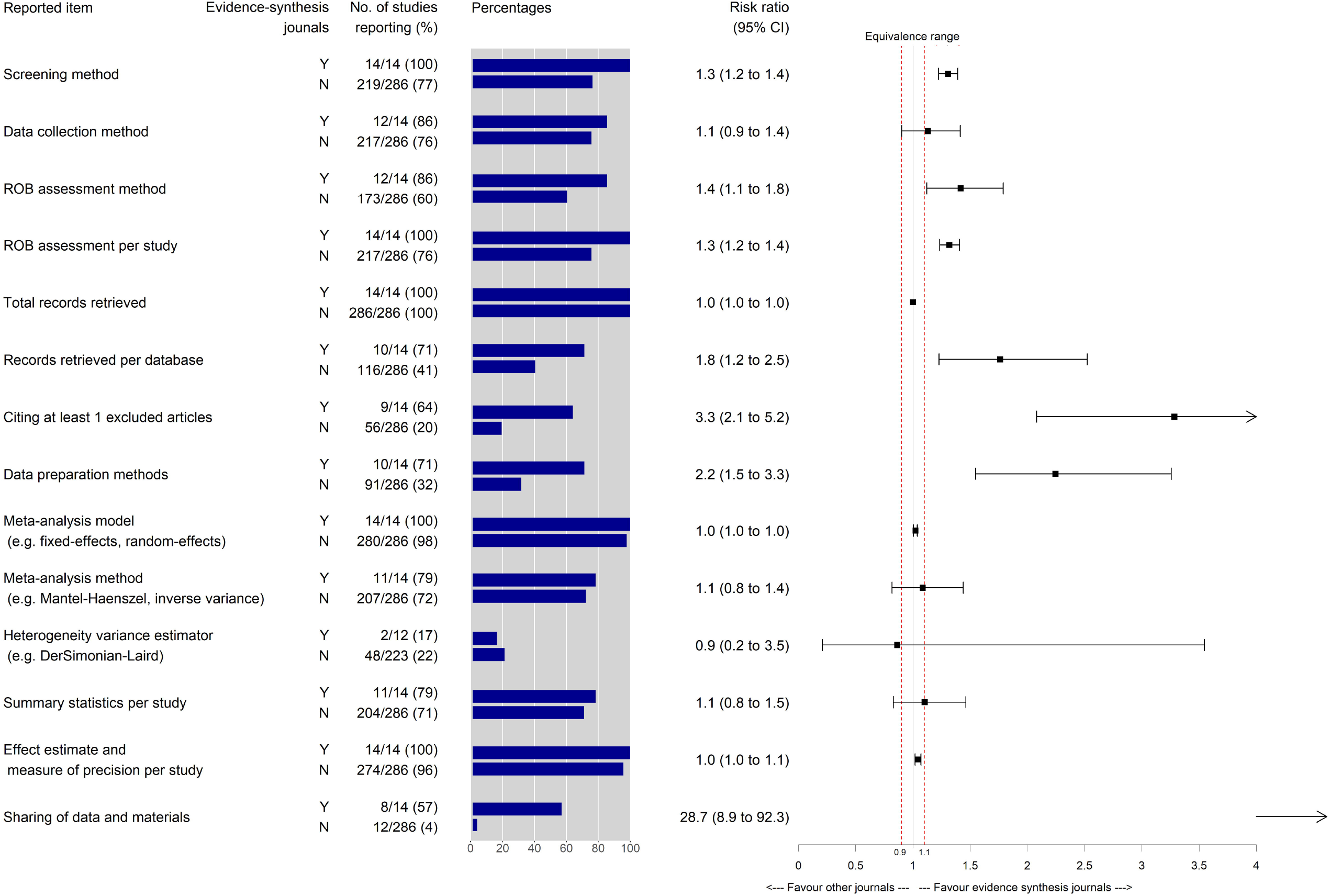
Relationship between journal type and reported items (ctn’d)

Systematic reviews published in a journal with a mandatory requirement for data sharing or declaration of data availability were more likely than reviews published elsewhere to share any data or materials (18% vs 2%) (Fig. 5). Similar findings were observed when comparing between journals with any data-sharing policy (mandatory or not) and journals without one (Supplemental data).

**Fig. 5.**
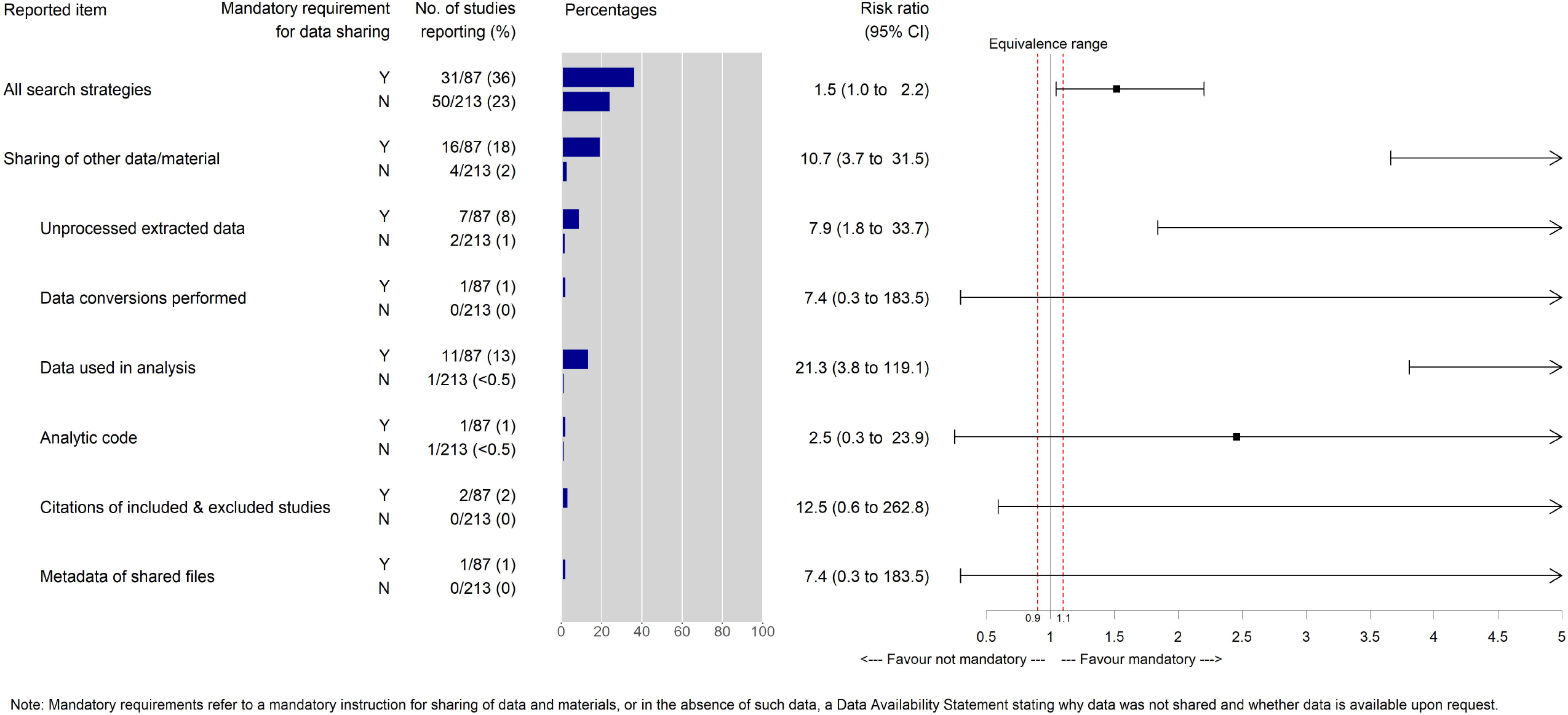
Relationship between journal’s data sharing requirements and reported items

## DISCUSSION

Findings from our examination of 300 randomly selected systematic reviews indexed in 2020 indicate suboptimal reporting of several items, such as the reporting of a review protocol (4%) or registration entry (38%), search strategy for all databases (27%), methods of handling data (e.g. imputing missing data, data conversions) (34%) and funding source for the review (72%). Other meta-research studies reported similar frequencies of reporting of review protocols (17%) [6], preregistration records (22%) [6], full search strategies for all databases (14%) [7], handling of missing data (25%) [4], and the funding source for the review (62%) [6]. Some discrepancies in these results can be attributed to differences in assessment criteria and the disciplines studied [34]. In our sample of reviews indexed in 2020, citation of reporting guidelines was common (82%), but there was no evidence that reviews that cited a guideline were reported more completely than reviews that did not, an observation shared by Wayant et al. [4]. We also reported a scarcity of data and code file sharing (7%), which is within the range of previously reported results (0.6%-11%) [4,8,35]. Journals’ open data policies were found to have positive impacts on the frequency of sharing certain types of review data and analytic code, which aligns with evaluations of other study designs [36,37].

### Strengths and limitations

Although this topic has been explored in other meta-research studies [2–8], our study offers several methodological advantages. Firstly, our assessment of reporting captured several recommended reporting items in the PRISMA 2020 statement [38] which have not previously been explored. Secondly, most previous meta-research studies on this topic used the 2009 PRISMA checklist to evaluate reporting [15], in which several reporting items comprise multiple elements (e.g. Item 10 reads “Describe method of data extraction from reports (such as piloted forms, independently, in duplicate) and any processes for obtaining and confirming data from investigators”). Simply recording “reported” for such an item does not clearly distinguish which elements in the item were actually reported. In contrast, the criteria we used to evaluate systematic reviews allowed for a more comprehensive and granular assessment of reporting in systematic reviews. Thirdly, our sample consists of systematic reviews published a few months before the PRISMA 2020 statement was released, and thus provides a useful benchmark for future meta-research studies to explore whether changes in reporting occurred after the release of PRISMA 2020. Fourthly, we searched several databases to identify eligible systematic reviews, and our sample was not limited to a specific topic or journal. Fifthly, our study captured not only the frequency of data sharing, but also the type of systematic review data, code and materials being shared. Lastly, we compared our 2020 sample with a 2014 sample that was retrieved and evaluated using the same criteria [9,10] thus minimising the impact of methodological variations.

Nonetheless, our study was not without limitations. We used web archives to determine the journal’s policies on data sharing prior to 1 November 2020 (i.e. just before the reviews in our sample were indexed in databases), but it was impossible to confirm with certainty the journal data policy that reviewers would have seen at the time of submission of their systematic review. As a cross-sectional study, our results should be viewed as generating hypotheses rather than proving a causal association. Some items were reported by fewer than 50 reviews, which caused uncertainty in interpreting their risk ratios. Despite intending to include systematic reviews of the effects of health, social, behavioural and educational interventions, the vast majority of reviews evaluated the effects of a health intervention. Therefore, our findings are less generalizable to systematic reviews of the other types of interventions. Lastly, our findings do not necessarily generalise to systematic reviews indexed in databases other than the ones we searched, or to systematic reviews written in languages other than English.

### On reporting of systematic reviews

There are several possible reasons why we observed few notable improvements in reporting between 2014 and 2020. Firstly, several items were already reported frequently in 2014 (e.g. reporting of competing interests, eligibility criteria, meta-analytic models, effect estimate for each study), leaving little opportunity for improvement. Secondly, some reporting items we examined have only been recommended for reporting recently (e.g. in the PRISMA 2020 statement published in March 2021) [38], such as search strategy for all databases or availability of data or analytic code. As such, authors of reviews in our study using older reporting guidelines might not have felt compelled to report these details in either 2014 or 2020.

Most systematic reviews in 2020 cited a reporting guideline, yet this was not clearly associated with improved reporting for any of the assessed items. This challenges the assumption that referencing a reporting guideline guarantees adherence to the guideline. In reality, other factors could have affected the authors’ decision not to report certain items. Firstly, authors might assume that reporting of methods used for one process implies that the same approach was used for another process. For example, we observed among our sample a tendency to report the reviewer arrangement only for screening stage, not the subsequent data collection or risk of bias assessment stages. Secondly, authors might incorrectly assume that the meta-analytic methods can always be deduced from knowing the packages and softwares used, or from reading the forest plot. This is not always the case [39], as different meta-analytic softwares have different options and default settings [40]. Thirdly, some items are difficult to report if the reviewer had not recorded relevant details during the conduct of the review (e.g. number of records excluded, data conversions performed). Fourthly, nearly all of the items reported in less than 50% of reviews, such as the interface used to search databases and meta-analysis method used, are recommended only in the explanation and elaboration document of the 2009 PRISMA statement, so these important elements might have been missed by authors using only the PRISMA checklist to guide reporting. In future, we recommend interviews be conducted with review authors to explore their understanding of reporting guidelines and identify challenges in reporting of reviews. Furthermore, interventions should be developed and evaluated to help improve reporting (such as a computer-based tool to break down the PRISMA reporting recommendations – both those appearing in the main checklist and those in the explanation and elaboration – into digestible steps for first-time reviewers [41,42]) and aid peer reviewers’ ability to detect incomplete reporting.

### On data sharing in systematic reviews

The low rate of data and code sharing can be attributed to several factors. Firstly, the issue of data sharing for systematic reviews has received relatively little attention until recently. A recommendation to report *whether* data, code and other materials are publicly available was only recommended in the PRISMA 2020 statement (published in March 2021), while our sample of systematic reviews were published prior to December 2020. Secondly, there has been a rise in percentage of non-Cochrane reviews between 2014-2020. Unlike Cochrane reviews, which are routinely published together with RevMan files containing meta-analysis data, non-Cochrane reviews are not always subjected to data sharing requirement. Third, there are motivational, educational, and technical barriers to data sharing that cannot be sufficiently addressed by data sharing policies, such as lack of technical expertise and time, lack of data management templates to facilitate sharing of review data, concerns about data ownership, fear of criticism and lack of career incentives [43,44]. Some studies have explored these barriers in general academia, but we are uncertain whether researchers in evidence synthesis face all of these barriers or even unidentified barriers unique to systematic reviews and meta-analyses. Future studies in the REPRISE project will explore systematic reviewers’ perspectives in order to address these questions [20].

Lastly, our findings also highlight the important role of supplementary files or public repositories for data sharing in systematic reviews. Web-based supplementary files and public repositories enable authors to share data and materials necessary to validate the review process while keeping the main article concise and relevant to lay readers [10]. For example, authors can outline in a separate file all database-specific search strategies (e.g. [45]), excluded studies at each stage of screening (e.g. [46]) and complete data for all meta-analyses (e.g. [47]). Data sharing via supplementary files or public repositories is an effective tool to improve reproducibility of systematic reviews and should be made a standard practice. Concerted efforts around data infrastructures, fair use guidelines and a supportive environment are required to make data sharing a standard practice [48–50].

## CONCLUSION

(www) Incomplete reporting of several recommended items in systematic reviews persists, even in reviews that claim to have followed a reporting guideline. Data sharing policies could be an effective strategy to promote sharing of systematic review data and materials.

### Copyright

The Corresponding Author has the right to grant on behalf of all authors and does grant on behalf of all authors, an exclusive licence (or non-exclusive for government employees) on a worldwide basis to the BMJ Publishing Group Ltd to permit this article (if accepted) to be published in BMJ editions and any other BMJPGL products and sublicences such use and exploit all subsidiary rights, as set out in our licence.

## Data Availability

All datasets and analytic code can be found on the Open Science Framework (DOI: 10.17605/OSF.IO/JSP9T).

https://osf.io/jsp9t/

## Acknowledgement

Not applicable

## Ethics approval

Not required as this study does not involve human participants or animal subjects

## Patient consent

Not required as this study does not involve human participants

## Transparency Declaration

The lead authors (PYN and MJP) affirm that the manuscript is an honest, accurate, and transparent account of the study being reported; that no important aspects of the study have been omitted; and that any discrepancies from the study as originally planned (and, if relevant, registered) have been explained.

## Data availability statement

All datasets and analytic code can be found on the Open Science Framework (DOI: <u>10.17605/OSF.IO/JSP9T</u>).

### Dissemination to participants and related patient and public communities

We plan to disseminate results of this study at national and international conferences, via seminars and workshops teaching systematic review methods (targeting clinicians, guideline developers, researchers and other stakeholders) and via Twitter.

## Contributors

The corresponding author attests that all listed authors meet authorship criteria and that no others meeting the criteria have been omitted.

P-YN: Data Curation, Formal Analysis, Investigation, Methodology, Writing – Original Draft Preparation RK, ZA, SM: Investigation, Writing – Review & Editing

JEM: Conceptualization, Supervision, Validation, Writing – Review & Editing

SEB, NRH, DGH, SK DM, SN, DN, PT, VAW: Writing – Review & Editing

MJP: Conceptualization, Funding Acquisition, Methodology, Supervision, Writing – Review & Editing

### Provenance and peer review

Not commissioned; externally peer reviewed

#### A CC BY licence is required

## Supplemental data

S1 Appendix: Deviations from protocol

S2 Appendix: Eligibility criteria for study inclusion

S3 Appendix: Search strategy

S4 Appendix: Data extraction form

S5 Appendix: List of journals and their policies

S6 Appendix: Comparison between reviews of health interventions in 2014 vs 2020

S7 Appendix: STROBE statement checklist

Figure S1: Sensitivity analysis – Frequency of reporting items between systematic reviews published in 2014 and 2020

Figure S2A: Sensitivity analysis – Relationship between citation of reporting guidelines and reported items

Figure S2B: Sensitivity analysis – Relationship between citation of reporting guidelines and reported items (ctn’d)

Figure S3: Relationship between journal’s presence of a data sharing policy and reported items

