## Supplemental data for "Changing patterns in reporting and sharing of review data in systematic reviews with meta-analysis of the effects of interventions: a meta-research study"

### TITLE PAGE

**Appendix S1. Deviations from the REPRISE project protocol**

| Original plan | Revised plan | Reason for modification |
|---|---|---|
| We will use risk ratios with 95% confidence intervals to examine differences in percentages of each indicator between reviews in different categories (e.g. 2020 versus 2014 reviews). | When the numerators were small (<5) in either group, or the outcome was very rare (<5%) in either group, rather than using risk ratios, we instead used penalised likelihood logistic regression and generated odds ratios. | Penalised likelihood logistic regression has been shown to improve estimation of the risk ratio and its confidence interval for rare events or unbalanced samples. The odds ratios from these models can be interpreted as risk ratios when the events are rare in both groups. |
| We will use risk ratios with 95% confidence intervals to examine differences in percentages of each indicator between reviews published in journals with versus without a mandatory data or code sharing policy | We will use risk ratios with 95% confidence intervals to examine differences in percentages of each indicator between reviews published in journals with versus without a policy that mandates either data sharing or declaration of data availability. | Too few journals had a policy that mandated data sharing, so we expanded this category to those journals which mandated inclusion of a data availability statement. |
| We did not plan to define an equivalence range to interpret risk ratio associations. | We defined an equivalence range for all comparisons as 0.9 to 1.1 – any risk ratio less than 0.9 or more than 1.1 was deemed as an important difference. | We set an equivalence range in this paper because there are so many statistical tests being undertaken. With all these tests, there is a temptation to focus on those that are statistically significant, even if unimportant. |
| We did not plan to conduct any sensitivity analyses. | We conducted two post-hoc sensitivity analyses, the first by excluding Cochrane reviews, and the second by excluding reviews on COVID-19. | We excluded Cochrane reviews because they are subjected to strict editorial processes to ensure adherence to methodological conduct and reporting standards (unlike systematic reviews published in other journals). We excluded reviews focusing on COVID-19 due to concerns about short publication turnarounds, which could have an impact on reporting quality. |

**Appendix S2. Eligibility criteria for study inclusion**

| |
|---|
| **Title and abstract screening** |
| 1. Based on the title/abstract, the article appears to be a completed systematic review with meta-analysis of studies (i.e. not a protocol for a systematic review, nor a systematic review without meta-analysis, nor an overview of systematic reviews, nor a scoping review); and |
| 2. The article appears to have compared the effects of at least two health, social, behavioural or educational interventions applied to humans. That is, any intervention designed to improve health ("a state of complete physical, mental and social well-being and not merely the absence of disease or infirmity"), promote social welfare and justice, change behaviour, or improve educational outcomes. |
| **Full-text screening** |
| 1. The article was written in English; and |
| 2. The review objective or question was clearly stated in the Introduction or Methods section; and |
| 3. The source(s) used to search for studies were reported; and |
| 4. Methods used to assess validity of the findings of the included studies (e.g. risk of bias assessment, methodological quality assessment) were reported in the Methods or Results section; and |
| 5. The review included randomised or non-randomised studies (or both) comparing the effects of at least two health, social, behavioural or educational interventions designed to improve health, promote social welfare and justice, change behaviour, or improve educational outcomes in humans. That is, any intervention designed to improve health ("a state of complete physical, mental and social well-being and not merely the absence of disease or infirmity"), promote social welfare and justice, change behaviour, or improve educational outcomes. |
| 6. References to all studies included in the review were cited, either in the standard reference list or in a supplementary file (full bibliographic citations of included studies must have been presented for the systematic review to be considered eligible). |
| 7. The review presented at least one pairwise meta-analysis of aggregate data, including at least two studies, using any effect measure (e.g. mean difference, risk ratio). Network meta-analyses were eligible if they included at least one direct (i.e. pairwise) comparison that fulfilled the abovementioned criterion. Meta-analyses of individual participant data were excluded. |

**Appendix S3. Search strategy**

All searches were run on December 3rd, 2020.

**PubMed**

(meta-analysis[PT] OR meta-analysis[TI] OR systematic[sb]) AND 2020/11/02:2020/12/02[EDAT]

**Science Citation Index Expanded (SCI-EXPANDED) and Social Sciences Citation Index (SSCI) via Web of Science**

(TI=meta-analysis OR AB=meta-analysis OR TS=meta-analysis OR TI="systematic review" OR AB="systematic review" OR TS="systematic review") AND LANGUAGE: (English) AND DOCUMENT TYPES: (Article OR Review)

Indexes=SCI-EXPANDED, SSCI; Timespan=Last 4 weeks

**Scopus via Elsevier**

TITLE("meta-analysis" OR "systematic review") AND ORIG-LOAD-DATE > 1604275200 AND ORIGLOAD-DATE < 1606867200 AND (LIMIT-TO(DOCTYPE, "ar") OR LIMIT-TO(DOCTYPE, "re"))

**Education Collection via ProQuest (added in 30 last days)**

MAINSUBJECT.EXACT.EXPLODE("Meta Analysis") OR ab(meta-analysis OR systematic review) OR ti(meta-analysis OR systematic review)

**Appendix S4. Data extraction form**

**Section 1. General characteristics**

| **Item** | **Response option** |
|---|---|
| Record ID: | Free text |
| Systematic review ID (surname year): | Free text |
| Enter initials of data collector: | Free text |
| Specify the title of the systematic review: | Free text |
| Specify the journal: | Free text |
| Specify the full name of the corresponding author of the systematic review: | Free text |
| Specify the email address of the corresponding author of the systematic review: | Free text |
| Specify the country of the corresponding author of the systematic review: | Free text |
| What was the source of funding for the systematic review? | O Non-profit (e.g. government, university/hospital/research institute, charitable foundation)<br>O For-profit (e.g. pharmaceutical company)<br>O Both non-profit and for-profit<br>O Unclear if the funder is for-profit or non-profit (please specify funder): ___________________<br>O Authors specified there was no funding for the systematic review<br>O Not reported |
| Did any of the systematic reviewers disclose financial conflicts of interest? | O Conflict of interest present (i.e. at least one systematic reviewer reported a financial conflict of interest of any type)<br>O No conflict of interest (i.e. all systematic reviewers stated they had no financial conflicts of interest)<br>O Missing (i.e. no disclosure statement) |
| Specify the total number of studies included in the systematic review: | Numeric |
| Specify the type of participant(s) indicated in the review objective(s) or question(s) (e.g. adults with lung cancer, children with asthma, university students) | Free text |
| What is the broad ICD-11 category investigated in this systematic review? | O Not applicable<br>O 01 Certain infectious or parasitic diseases<br>O 02 Neoplasms<br>O 03 Diseases of the blood or blood-forming organs |

| | |
|---|---|
| | O 04 Diseases of the immune system<br>O 05 Endocrine, nutritional or metabolic diseases<br>O 06 Mental, behavioural or neurodevelopmental disorders<br>O 07 Sleep-wake disorders<br>O 08 Diseases of the nervous system<br>O 09 Diseases of the visual system<br>O 10 Diseases of the ear or mastoid process<br>O 11 Diseases of the circulatory system<br>O 12 Diseases of the respiratory system<br>O 13 Diseases of the digestive system<br>O 14 Diseases of the skin<br>O 15 Diseases of the musculoskeletal system or connective tissue<br>O 16 Diseases of the genitourinary system<br>O 17 Conditions related to sexual health<br>O 18 Pregnancy, childbirth or the puerperium<br>O 19 Certain conditions originating in the perinatal period<br>O 20 Developmental anomalies<br>O 21 Symptoms, signs or clinical findings, not elsewhere classified<br>O 22 Injury, poisoning or certain other consequences of external causes<br>O 23 External causes of morbidity or mortality<br>O 24 Factors influencing health status or contact with health services |
| Specify the type of intervention(s) indicated in the review objective(s) or question(s) (e.g. inhaled corticosteroids, provision of charity or welfare, use of bystander programs, reduction in class size) [Select all that apply]: | ☐ Health (i.e. any intervention designed to improve “health” defined as a state of complete physical, mental and social well-being and not merely the absence of disease or infirmity)<br>☐ Behavioural (i.e. any intervention designed to increase useful behaviours and reduce or eliminate harmful behaviours, such as modelling, prompting, reinforcement, or shaping behaviours)<br>☐ Educational (i.e. any intervention designed to provide students with the support needed to acquire the skills being taught by an educational system)<br>☐ Social (i.e. any intervention of a government or an organization in social affairs, such as provision of charity or social welfare as a means to alleviate social and economic problems of people facing financial difficulties; provision of safety regulations for employment and products; delivery of food aid, food bank or recovery missions to regions or countries negatively affected by an event; securing workers’ rights through gradualism in institutions, private education and unions) |
| Is/are the intervention intervention(s) indicated in the review objective(s) or question(s) pharmacological or non-pharmacological? | O Pharmacological |

| | |
|---|---|
| | O Non-pharmacological (i.e. any intervention that does not involve drugs, e.g. device, vitamin, diet, psychotherapy, physical therapy, welfare, organization change, education)<br>O Both pharmacological and non-pharmacological |
| Notes | Free text |

**Section 2. Transparency characteristics**

| Item | Response option |
|---|---|
| Did the authors state that the systematic review was reported in accordance with a reporting guideline (e.g. PRISMA, MOOSE, MECIR, MARS, ROSES)? | O Yes<br>O No |
| Is a protocol or registration record for the systematic review cited within the review? | O Both a protocol and registration record are cited<br>O Only a protocol is cited<br>O Only a registration record is cited<br>O Neither are cited |
| Did the authors specify any eligibility criteria for the following components of the review question [Select all that apply]? | ☐ Participants<br>☐ Interventions/exposures<br>☐ Comparators<br>☐ Outcomes |
| Did the authors make a statement regarding eligibility of studies based on study design? | O Yes<br>O No |
| [If "Yes" to the previous question] Specify the type of study design(s) eligible for inclusion in the systematic review: | O Randomised trial<br>O Non-randomised study (e.g. cohort study, case-control study)<br>O Both randomised trials and non-randomised studies |
| How many bibliographic databases were searched? [note: trials registers (e.g. WHO ICTRP, ClinicalTrials.gov) are not applicable to this question] | Numeric |
| Copy and paste verbatim the list of databases searched: | Free text |
| Was the interface or platform through which the bibliographic database was searched reported for any database (e.g. did the authors specify that MEDLINE was searched via Ovid, or that CINAHL was searched via EBSCOhost)? | O Fully – interface or platform reported for all databases<br>O Partially – interface or platform reported for only some databases<br>O No – not reported for any database |

| | |
|---|---|
| Were the years of coverage reported for any bibliographic database searched? | O Fully – both start and end exact dates were reported [Select this option if both dates are reported, or if reviewers placed no limit/restriction on the start date and reported the date the search was conducted]<br>O Partially – both start and end dates were reported, but not as exact dates (e.g. MM/YY or WW/MM)<br>O Partially – only the start date or only the end date (e.g. "we searched all databases on 23 May 2019" or "we searched all databases from inception to current")<br>O Not reported |
| Did the authors specify the exact date when each bibliographic database was last searched? | O Yes<br>O No |
| Was the full search strategy or search terms used in the bibliographic databases reported? | O Yes – Full Boolean search logic was reported for all databases<br>O Yes – Full Boolean search logic was reported for only some databases<br>O Yes – Only main index terms (e.g. MeSH terms) used in the search strategy were reported<br>O Yes – Only free text terms used in the search strategy were reported<br>O No – Authors did not report any search strategy or search terms |
| Did the authors report searching a trials register (e.g. ClinicalTrials.gov, WHO International Clinical Trials Registry Platform)? | O Yes<br>O No |
| Did the authors report searching any other electronic source (e.g. Google Scholar, organisation websites, journal websites)? | O Yes<br>O No |
| If yes to the previous question, was the search logic that was used to search each of these sources reported? | O Fully – Search logic reported for all these sources<br>O Partially – Search logic reported for only some of these sources<br>O No – Search logic not reported for any of these sources |
| What method of study selection did the authors report using? | O All identified studies screened by at least two authors<br>O All identified studies screened by one author, with a sample screened by another<br>O At least two authors were involved in either titles/abstracts or full-text screening, but authors did not specify for the remaining step |

| | |
|---|---|
| | O All identified studies screened by only one author<br>O Different methods were applied for titles/abstracts and full-text screening<br>O Not reported/unclear |
| What method of data extraction did the authors report using? | O All data extracted by at least two authors<br>O All data extracted by one author, with verification by another<br>O All data extracted by only one author<br>O Other (please specify): ___________________<br>O Not reported/Unclear |
| What method of risk of bias (or quality) assessment did the authors report using? | O All included studies assessed by at least two authors<br>O All included studies assessed by one author, with verification by another<br>O All included studies assessed by only one author<br>O Other (please specify): ___________________<br>O Not reported/Unclear |
| Did the authors present data on risk of bias (or quality) for each item on a per-study basis (e.g. presented a table or figure displaying how each study was rated on each of the risk of bias (or quality) items)? | O Yes<br>O No |
| Which statistical software was used to perform meta-analyses [Select all that apply]? | ☐ R<br>☐ Stata<br>☐ RevMan<br>☐ Comprehensive Meta-Analysis (CMA)<br>☐ SAS<br>☐ SPSS<br>☐ Other (please specify)<br>☐ Not reported |
| Did the authors report which statistical package(s) were used (e.g. metan in Stata, metafor in R) and the version number of the package(s)? | O Both the statistical package(s) and software version number(s) were reported<br>O Only the statistical package(s) were reported |

| | |
|---|---|
| | O Only the software version number(s) were reported<br>O Neither were reported |
| Did the authors report the total number of records identified through all searches? | O Yes<br>O No |
| Did the authors report the number of records identified through each database/source (e.g. indicated that the search of MEDLINE identified 1000 records, Embase identified 1300 records, CENTRAL identified 750 records, etc.)? | O Yes<br>O No |
| Did the authors cite any full text articles that were screened and excluded from the review? | O Yes<br>O No |
| **Answer the following questions for only one meta-analysis per review (the index meta-analysis)**<br>**Select first reported meta-analysis. Note that the first meta-analysis may be identified from the Abstract or Results section of the review, depending on where it is first reported in the publication.** | |
| What is the outcome domain (e.g. all-cause mortality, anxiety)? | Text |
| How many studies were included in the meta-analysis? | Numeric |
| What was the effect measure for the meta-analysis? | O Risk ratio<br>O Odds ratio<br>O Hazard ratio<br>O Risk difference<br>O Mean difference<br>O Standardized mean difference<br>O Other (please specify): ____________________<br>O Not reported |

| | |
|---|---|
| Did the authors describe any methods required to prepare the data for the meta-analysis, such as handling of missing summary statistics, or data conversions? | O Yes<br>O No |
| Did the authors specify the meta-analysis model used (e.g. fixed-effect, random-effects)? [Note: such information may appear in the Methods section or on the forest plot] | O Yes<br>O No |
| Did the authors specify the meta-analysis method used (e.g. Mantel-Haenszel, inverse-variance)? [Note: such information may appear in the Methods section or on the forest plot] | O Yes<br>O No |
| Did the authors specify the between-study (heterogeneity) variance estimator used (e.g. DerSimonian and Laird, restricted maximum likelihood (REML)? [Note: Such information may appear in the Methods section or on the forest plot. Only answer as “Yes” if the estimator was reported explicitly. Answer as “No” if the estimator was not reported explicitly, even if you can guess the estimator from the software used (e.g. because the software implements only one option)] | O Yes<br>O No<br>O Not applicable – not a random-effects meta-analysis |
| Were summary statistics for each included study reported in a table, figure, or text? By “summary statistics”, we mean the mean, standard deviation and sample size for continuous outcomes, or the number of events and sample size for binary outcomes. | O Yes<br>O No |
| Were effect estimates and measures of precision of each included study reported in a table, figure, or text? [Notes: By “effect estimate”, we mean a point estimate of the intervention effect, such as a mean difference for continuous outcomes, or a risk ratio, odds ratio or risk difference for binary outcomes. By “measure of precision”, we mean a standard error or confidence interval.] | O Yes<br>O No |
| Is there a data or code availability statement in the paper? | O Yes<br>O No |

| | |
|---|---|
| If yes to the previous question, copy and paste the data and code statement(s) verbatim: | Free text |
| Have the authors made any of the following publicly accessible, either as a supplementary file or file uploaded to a general-purpose repository (e.g. Open Science Framework, Zenodo, GitHub) or institutional repository? [Select all that apply. Only select each if you are able to locate the relevant material, after searching all supplementary materials or links to shared materials] | ☐ Template data collection form(s)<br>☐ File(s) containing (unprocessed) data extracted from included studies<br>☐ File(s) indicating any necessary data conversions performed<br>☐ File(s) containing data used in all analyses (e.g. Microsoft Excel or CSV spreadsheet, or RevMan file containing all study effect estimates included in meta-analyses)<br>☐ Analytic code used to generate results (i.e. the sequence of commands used within a software package to manage and analyse data)<br>☐ File(s) containing citations of all records that were screened and excluded<br>☐ Metadata which describes the contents of the shared file(s) to aid interpretation and reuse (e.g. a file with complete descriptions of variable names, or README files describing each file shared) |
| If yes to any of the materials selected, how was the material shared publicly? [Select all that apply] | O Uploaded as a supplementary file on the journal website<br>O Uploaded to a general-purpose open-access repository (e.g. Open Science Framework, Zenodo, GitHub) (please specify)<br>O Uploaded to an institutional repository<br>O Uploaded to my personal website<br>O Other (please specify): ____________________ |
| If yes to any of the materials selected, are any of the data file(s) associated with a persistent identifier (e.g. DOI)? | O Yes<br>O No<br>O Unsure |
| If yes to any of the materials selected, was a license applied to any of the data file(s) (e.g. CC BY, CC BY-NC)? | O Yes<br>O No<br>O Unsure |
| Notes | Free text |

**Section 3. Journal characteristics.**

| | |
|---|---|
| 1. Journal name (Abbreviated) | Free text |
| 2. Number of studies published under this journal | Numeric |
| 3. Does this journal only publish evidence syntheses (reviews, meta-analyses and their protocols)? | O Yes<br>O No |
| **"Instruction for Authors" Section** | |
| 4. Does this journal indicate a data or code sharing policy (or both) under instructions to authors or submission guidelines? | O Yes – The journal encourages or mandates that authors of all research articles (or systematic reviews in particular) share their data or analytic code when they submit an article, as specified in the journal instructions to authors or submission guidelines<br>O No - No such policy can be located in the journal instructions to authors or submission guidelines |
| 5. (If yes to Q4) Copy policy text verbatim | Free text |
| 6. (If yes to Q4) What is the expectation for authors regarding sharing of data and materials (including analytic code)? | O Encouraged: both sharing of data and/or analytic code and the Data Availability Statement are not mandatory for publication by the journal<br>O Expected: sharing data and/or analytic code is not mandatory for publication of systematic reviews, but a Data Availability Statement, which contains links to shared data or reasons for not sharing data, must be provided.<br>O Mandatory: sharing data and/or analytic code is a condition of publication of systematic reviews by the journal; |

#### Appendix S5. List of journals and their policies

| Journal | Evidence synthesis journal | Data policy present | Type of data policy | Publisher | Earliest recorded date of journal's data policy * | Earliest recorded date of publisher's data policy ** |
|---|---|---|---|---|---|---|
| Acad Emerg Med | No | Yes | Encouraged | Wiley OBO SAEM | 14-Sep-21 | 14-Sep-17 |
| Accid Anal Prev | No | Yes | Encouraged | Elsevier | 8-Aug-20 | - |
| Acta Anaesthesiol Scand | No | Yes | Encouraged | Wiley-Blackwell | 30-May-21 | 14-Sep-17 |
| Acta Diabetol | No | Yes | Encouraged | Springer | 28-Jul-20 | - |
| Acta Neurol Scand | No | Yes | Expected | Wiley-Blackwell | 27-May-19 | - |
| Acta Ophthalmol | No | Yes | Encouraged | Wiley-Blackwell | 29-Nov-19 | - |
| Acta Psychiatr Scand | No | Yes | Expected | Wiley-Blackwell | 5-May-19 | - |
| Addiction | No | Yes | Encouraged | Wiley-Blackwell | 4-May-19 | - |
| Adv Ther | No | No | - | Springer | - | - |
| Adv Nutr (Bethesda, Md) | No | No | - | ASN | - | - |
| Aids | No | No | - | Lippincott W&W | - | - |
| Aliment Pharmacol Ther | No | Yes | Expected | Wiley-Blackwell | 5-May-19 | - |
| Am Heart J | No | Yes | Encouraged | Elsevier | 30-Jan-19 | - |
| Am J Cardiol | No | Yes | Encouraged | Elsevier | 26-Jan-20 | - |
| Am J Mens Health | No | No | - | SAGE | - | - |
| Am J Sports Med | No | No | - | SAGE | - | - |
| Ann Palliat Med | No | No | - | Mary Ann Liebert | - | - |
| Ann Transl Med | No | No | - | AME Publishing | - | - |
| Ann Surg | No | No | - | Lippincott W&W | - | - |
| Ann Roy Coll Surg | No | No | - | Royal College of Surgeons of England | - | - |
| Antioxidants (Basel) | No | Yes | Mandatory | MDPI | 12-May-21 | 10-Oct-20 |
| Arch Public Health | No | Yes | Expected | BMC | n/a | 12-Jun-16 |
| Arthritis Rheumatol | No | Yes | Mandatory | Wiley & Sons | 21-Oct-20 | - |
| Arthrosc Sports Med Rehabil | No | Yes | Encouraged | Elsevier | 1-Aug-20 | - |
| Attach Hum Dev | No | Yes | Expected | Taylor & Francis | n/a | 13-Feb-18 |
| Aust Dent J | No | Yes | Encouraged | Wiley-Blackwell | 8-Mar-22 | 14-Sep-17 |
| Aust J Prim Health | No | Yes | Expected | CSIRO | 4-Jul-22 | 4-Sep-17 |
| Autoimmun Rev | Yes | No | - | Elsevier | - | - |
| Biomed Res Int | No | No | - | Hindawi | - | - |
| Bioscience Rep | No | Yes | Expected | Portland Press | 28-Sep-20 | - |
| Birth Defects Res | No | Yes | Expected | Wiley & Sons | 17-Jul-19 | - |
| BJOG | No | Yes | Expected | Wiley-Blackwell | 19-Jun-20 | - |
| BJS Open | No | Yes | Encouraged | Oxford Uni | 5-Apr-22 | Early 2020 |
| Blood Purif | No | Yes | Encouraged | Karger AG | 14-Sep-20 | - |
| BMC Musculoskelet Disord | No | Yes | Expected | BMC | n/a | 12-Jun-16 |
| BMC Psychiatry | No | Yes | Expected | BMC | n/a | 12-Jun-16 |
| BMC Urol | No | Yes | Expected | BMC | n/a | 12-Jun-16 |
| BMJ | No | Yes | Expected | BMJ | 11-Jan-18 | - |
| BMJ Open | No | Yes | Expected | BMJ | 30-Jan-19 | - |
| BMJ Open Sport Exerc Med | No | Yes | Expected | BMJ | 15-Dec-19 | - |
| BMJ Support Palliat Care | No | Yes | Expected | BMJ | 7-Jan-19 | - |

| Journal | Evidence synthesis journal | Data policy present | Type of data policy | Publisher | Earliest recorded date of journal's data policy * | Earliest recorded date of publisher's data policy ** |
|---|---|---|---|---|---|---|
| Bone Rep | No | Yes | Encouraged | Elsevier | 24-Dec-19 | - |
| Br J Anaesth | No | Yes | Encouraged | Elsevier | 24-Dec-19 | - |
| Br J Sports Med | No | Yes | Expected | BMJ | 11-Jan-18 | - |
| Braz Oral Res | No | No | - | Sao Paolo Uni | - | - |
| Can J Cardiol | No | Yes | Encouraged | Elsevier | 25-Sep-20 | - |
| Can Respir J | No | No | - | Hindawi | - | - |
| Cancer | No | Yes | Encouraged | Wiley & Sons | 22-Sep-20 | - |
| Cancers | No | Yes | Mandatory | MDPI | 25-Nov-20 | 10-Oct-20 |
| Cardiol J | No | Yes | Encouraged | Via Medica | 16-Jul-20 | - |
| Cardiovasc Drugs Ther | No | Yes | Encouraged | Springer | 30-Jul-21 | 5-Jul-16 |
| Cardiovasc Interv Ther | No | No | - | Springer | - | - |
| Cardiovasc Revasc Med | No | Yes | Encouraged | Elsevier | 10-Jun-21 | 4-Sep-17 |
| Catheter Cardiovasc Interv | No | No | - | Wiley-Liss | - | - |
| Clin Cardiol | No | No | - | Wiley-Blackwell | - | - |
| Clin Infect Dis | No | Yes | Encouraged | Oxford Uni | 22-May-22 | Early 2020 |
| Clin Microbiol Infect | No | No | - | Elsevier | - | - |
| Clin Nutr | No | Yes | Encouraged | Elsevier | 7-Sep-20 | - |
| Clin Nutr ESPEN | No | Yes | Encouraged | Elsevier | 20-Sep-20 | - |
| Clin Oral Investig | No | Yes | Encouraged | Springer | 24-Sep-20 | - |
| Clin Rehabil | No | No | - | SAGE | - | - |
| Clin Transl Oncol | No | Yes | Encouraged | Springer | 28-Nov-21 | 5-Jul-16 |
| Clin Neuropharmacol | No | No | - | Lippincott W&W | - | - |
| Cochrane Database Syst Rev | Yes | Yes | Mandatory | Wiley & Sons | 1-Apr-14 | - |
| Complement Ther Clin Pract | No | Yes | Encouraged | Elsevier | 22-Oct-20 | - |
| Complement Ther Med | No | Yes | Encouraged | Elsevier | 7-Sep-20 | - |
| Crit Rev Oncol Hematol | Yes | Yes | Encouraged | Elsevier | 4-Apr-20 | - |
| Dan Med J | No | No | - | Almindelige Danske Laegeforening | - | - |
| Dermatol Ther | No | Yes | Expected | Wiley-Blackwell | 18-Oct-20 | - |
| Diabetes Metab Syndr: Clin Res Rev | No | Yes | Encouraged | Elsevier | n/a | 4-Sep-17 |
| Diabetes Ther | No | No | - | Springer | - | - |
| Diagn Interv Radiol | No | No | - | Turkish Society of Radiology | - | - |
| Disabil Rehabil | No | Yes | Encouraged | Taylor & Francis | n/a | 13-Feb-18 |
| Drug Saf | No | Yes | Encouraged | Springer | 9-Aug-20 | - |
| EClinicalMedicine | No | Yes | Expected | Lancet | 21-Sep-20 | - |
| Eur Heart J | No | Yes | Expected | Oxford Uni | 29-Mar-20 | - |
| Eur Heart J Cardiovasc Pharmacother | No | Yes | Expected | Oxford Uni | 15-May-20 | - |
| Eur J Anaesth | No | No | - | Lippincott W&W | - | - |
| Eur J Cancer | No | Yes | Encouraged | Elsevier | 14-Aug-20 | - |
| Eur J Clin Microbiol Infect Dis | No | No | - | Springer | - | - |
| Eur J Hosp Pharm | No | Yes | Expected | European Association of Hospital Pharmacists | 4-Jan-19 | - |
| Eur J Nutr | No | Yes | Encouraged | Springer | 19-May-20 | - |
| Eur J Ophthalmol | No | No | - | Wichtig | - | - |

| Journal | Evidence synthesis journal | Data policy present | Type of data policy | Publisher | Earliest recorded date of journal's data policy * | Earliest recorded date of publisher's data policy ** |
|---|---|---|---|---|---|---|
| Eur J Pediatr | No | Yes | Encouraged | Springer | 10-Aug-20 | - |
| Eur J Pediatr Surg | No | No | - | Thieme | - | - |
| Eur Radiol | No | No | - | Springer | - | - |
| Eur Rev Med Pharmacol Sci | Yes | No | - | Verduci Editore | - | - |
| Eur Spine J | No | Yes | Encouraged | Springer | 30-Aug-20 | - |
| Eur J Integr Med | No | Yes | Encouraged | Elsevier | 7-Sep-20 | - |
| Evid Based Complement Alternat Med | No | No | - | Hindawi | - | - |
| Exp Gerontol | No | Yes | Encouraged | Elsevier | 7-Sep-20 | - |
| Food Funct | No | No | - | Royal Society of Chemistry | - | - |
| Foot Ankle Surg | No | Yes | Encouraged | Elsevier | 5-Aug-20 | - |
| Front Oncol | No | No | - | Frontiers Media | - | - |
| Front Pharmacol | No | No | - | Frontiers Media | - | - |
| Front Psychiatry | No | No | - | Frontiers Media | - | - |
| Front Surg | No | No | - | Frontiers Media | - | - |
| Gastroenterol Hepatol | No | Yes | Encouraged | Elsevier | 26-Aug-16 | - |
| Gen Thorac Cardiovasc Surg | No | No | - | Springer | - | - |
| Gerodontology | No | Yes | Encouraged | Wiley-Blackwell | 8-Mar-22 | 14-Sep-17 |
| Hepatology | No | Yes | Mandatory | Wiley & Sons | n/a | 14-Sep-17 |
| Horm Mol Biol Clin Investig | No | No | - | de Gruyter | - | - |
| Hum Reprod Update | No | Yes | Expected | Oxford Uni | 17-May-20 | - |
| Indian J Med Microbiol | No | Yes | Encouraged | Elsevier | 29-Nov-21 | 4-Sep-17 |
| Indian J Ophthalmol | No | No | - | Wolters Kluwer | - | - |
| Indian J Surg | No | Yes | Encouraged | Springer | 7-Aug-20 | - |
| Injury | No | Yes | Encouraged | Elsevier | 8-Aug-20 | - |
| Int Braz J Urol | No | No | - | Brazilian Society of Urology | - | - |
| Int Breastfeed J | No | Yes | Expected | BMC | n/a | 12-Jun-16 |
| Int J Clin Pract | No | Yes | Encouraged | Wiley-Blackwell / Hindawi (22 Sep 2021 onwards) | 30-Mar-22 | 14-Sep-17 |
| Int J Endocrinol | No | No | - | Hindawi | - | - |
| Int J Environ Res Public Health | No | Yes | Mandatory | MDPI | 29-Oct-20 | 10-Oct-20 |
| Int J Paediatr Dent | No | Yes | Encouraged | Wiley-Blackwell | 8-Mar-22 | 14-Sep-17 |
| Int J Rehabil Res | No | No | - | Lippincott W&W | - | - |
| Int J Vitam Nutr Res | No | No | - | Hogrefe | - | - |
| Int Wound J | No | Yes | Encouraged | Wiley-Blackwell | 18-Aug-21 | 14-Sep-17 |
| Integr Cancer Ther | No | Yes | Mandatory | SAGE | 24-Apr-20 | - |
| Integr Med Res | No | Yes | Encouraged | Elsevier | 15-Jan-19 | - |
| Iran J Microbiol | No | No | - | Teheran University of Medical Sciences | - | - |
| J Adolesc | No | Yes | Encouraged | Elsevier / Wiley (as of 1 Jan 2022) | 15-Oct-18 | - |
| J Affect Disord | No | Yes | Encouraged | Elsevier | 22-Oct-20 | - |
| J Altern Complement Med | No | Yes | Encouraged | Mary Ann Liebert | 21-Sep-20 | - |
| J Am Med Dir Assoc | No | Yes | Encouraged | Elsevier | 24-Oct-19 | - |
| J Arthroplasty | No | Yes | Encouraged | Elsevier | 3-Feb-21 | 4-Sep-17 |
| J Bodyw Mov Ther | No | No | - | Churchill Livingstone | - | - |

| Journal | Evidence synthesis journal | Data policy present | Type of data policy | Publisher | Earliest recorded date of journal's data policy * | Earliest recorded date of publisher's data policy ** |
|---|---|---|---|---|---|---|
| J Cardiovasc Pharmacol | No | No | - | Lippincott W&W | - | - |
| J Clin Med | No | Yes | Mandatory | MDPI | 11-Nov-20 | 10-Oct-20 |
| J Clin Neurosci | No | Yes | Encouraged | Elsevier | 29-Aug-17 | - |
| J Clin Pharm Ther | No | No | - | Wiley-Blackwell | - | - |
| J Coll Physicians Surg Pak | No | No | - | College of Physicians and Surgeons Pakistan | - | - |
| J Consult Clin Psychol | No | Yes | Expected | APA | 8-Aug-17 | - |
| J Craniofac Surg | No | No | - | Lippincott W&W | - | - |
| J Crit Care | No | Yes | Encouraged | Elsevier | 7-Sep-20 | - |
| J Dent Anesth Pain Med | No | No | - | Korean Dental Society of Anesthesiology | - | - |
| J Diabetes Investig | No | No | - | Blackwell Asia | - | - |
| J Emerg Med | No | Yes | Encouraged | Elsevier | 4-Sep-18 | - |
| J Food Biochem | No | Yes | Expected | Wiley-Blackwell | n/a | 14-Sep-17 |
| J Formos Med Assoc | No | Yes | Encouraged | Excerpta Medica | 11-Nov-19 | - |
| J Gastroenterol Hepatol | No | Yes | Encouraged | Wiley-Blackwell | 4-May-19 | - |
| J Gynecol Oncol | No | No | - | Korean Society of Gynecologic Oncology and Colposcopy | - | - |
| J Hepato-Biliary-Pancreat Sci | No | Yes | Encouraged | Wiley-Blackwell | n/a | 14-Sep-17 |
| J Herb Med | No | Yes | Encouraged | Elsevier | 11-Aug-20 | - |
| J Int AIDS Soc | No | Yes | Expected | Wiley & Sons | 25-Jul-20 | - |
| J Int Med Res | No | Yes | Mandatory | SAGE | 26-Jun-20 | - |
| J Intensive Care | No | Yes | Expected | BMC | n/a | 12-Jun-16 |
| J Interv Cardiol | No | No | - | Wiley-Blackwell | - | - |
| J Invest Surg | No | Yes | Encouraged | Taylor & Francis | 15-Jun-22 | 13-Feb-18 |
| J Matern Fetal Neonatal Med | No | Yes | Encouraged | Taylor & Francis | n/a | 13-Feb-18 |
| J Med Internet Res | No | Yes | Encouraged | JMIR | 5-Aug-20 | - |
| J Minim Invasive Gynecol | No | Yes | Encouraged | Elsevier | 7-Sep-20 | - |
| J Obstet Gynaecol | No | Yes | Encouraged | Taylor & Francis | n/a | 13-Feb-18 |
| J Ophthalmol | No | No | - | Hindawi | - | - |
| J Oral Biol Craniofac Res | No | Yes | Encouraged | Elsevier | 5-Sep-20 | - |
| J Oral Implantol | No | No | - | Allen Press | - | - |
| J Orthop Surg Res | No | Yes | Expected | BMC | n/a | 12-Jun-16 |
| J Pediatr Urol | No | Yes | Encouraged | Elsevier | 21-Aug-20 | - |
| J Pharm Sci | No | No | - | Wiley & Sons | - | - |
| J Prosthet Dent | No | Yes | Encouraged | Elsevier | 23-Aug-22 | 4-Sep-17 |
| J Rehabil Med | No | No | - | Foundation for Rehabilitation Information | - | - |
| J Shoulder Elbow Surg | No | No | - | Mosby | - | - |
| J Surg Res | No | Yes | Encouraged | Elsevier | 9-Mar-22 | 4-Sep-17 |
| J Tradit Chin Med | No | No | - | Journal of Traditional Chinese Medicine | - | - |
| J Vasc Surg | No | No | - | Mosby | - | - |
| J Wound Care | No | No | - | MA Healthcare | - | - |
| Jama | No | No | - | AMA | - | - |
| JAMA Psychiatry | No | No | - | AMA | - | - |

| Journal | Evidence synthesis journal | Data policy present | Type of data policy | Publisher | Earliest recorded date of journal's data policy * | Earliest recorded date of publisher's data policy ** |
|---|---|---|---|---|---|---|
| JMIR Serious Games | No | No | - | JMIR | - | - |
| J Clin Anesth | No | Yes | Encouraged | Elsevier | 27-Jul-20 | - |
| J Endourol | No | No | - | Mary Ann Liebert | - | - |
| J Hosp Infect | No | Yes | Encouraged | Elsevier | 8-Mar-21 | 4-Sep-17 |
| Knee Surg Sports Traumatol Arthrosc | No | Yes | Encouraged | Springer | n/a | 5-Jul-16 |
| Korean J Intern Med | No | No | - | Korean Association of Internal Medicine | - | - |
| Life (Basel) | No | Yes | Mandatory | MDPI | 21-May-21 | 10-Oct-20 |
| Medicine (Baltimore) | No | Yes | Expected | Lippincott W&W | 8-Mar-20 | - |
| Minerva Anestesiol | No | No | - | Minerva Medica | - | - |
| Neurochirurgie | No | Yes | Encouraged | Elsevier | 9-Sep-20 | - |
| Neurology | No | Yes | Mandatory | Lippincott W&W | 26-Jul-20 | - |
| Nurs Crit Care | No | Yes | Encouraged | Wiley-Blackwell | 9-Jun-21 | 14-Sep-17 |
| Nutrients | No | Yes | Mandatory | MDPI | 25-Oct-20 | 10-Oct-20 |
| Obes Rev | Yes | Yes | Encouraged | Wiley-Blackwell | 6-May-19 | - |
| Obesity (Silver Spring) | No | No | - | Wiley-Blackwell | - | - |
| Oper Dent | No | No | - | Indiana University School of Dentistry | - | - |
| Oral Health Prev Dent | No | No | - | Quintessence Publishing | - | - |
| Orthop Traumatol Surg Res | No | Yes | Encouraged | Elsevier | 7-Aug-20 | - |
| PACE-Pacing Clin Electrophysiol | No | No | - | Wiley-Blackwell | - | - |
| Pediatr Surg Int | No | Yes | Encouraged | Springer | 26-Nov-20 | 5-Jul-16 |
| Pediatrics | No | No | - | American Academy of Pediatrics | - | - |
| Pharmacol Res | No | Yes | Encouraged | Elsevier | 11-Aug-20 | - |
| Phys Ther | No | Yes | Encouraged | Oxford Uni | 29-Mar-20 | - |
| Physiother Theory Pract | No | Yes | Encouraged | Taylor & Francis | 23-Feb-22 | 13-Feb-18 |
| Phytother Res | No | Yes | Expected | Wiley & Sons | 17-Aug-21 | 14-Sep-17 |
| PLoS One | No | Yes | Mandatory | PLOS | 8-May-15 | - |
| Pol Intern Med | No | No | - | Medycyna Praktyczna | - | - |
| Porto Biomed J | No | No | - | Elsevier | - | - |
| Postgrad Med J | No | Yes | Expected | BMJ | 11-Jan-18 | - |
| Prog Neuropsychopharmacol Biol Psychiatry | No | Yes | Encouraged | Elsevier | 22-Oct-20 | - |
| Psychiatry Res | No | Yes | Encouraged | Elsevier | 21-Nov-18 | - |
| Psychol Med | No | No | - | Cambridge Uni | - | - |
| Psychol Sport Exerc | No | No | - | Human Kinetics | - | - |
| Reg Anesth Pain Med | No | Yes | Expected | BMJ | 11-Jan-18 | - |
| Reprod Biomed Online | No | Yes | Encouraged | Elsevier | 30-Oct-20 | - |
| Scand J Gastroenterol | No | Yes | Encouraged | Taylor & Francis | n/a | 13-Feb-18 |
| Schizophr Res | No | No | - | Elsevier | - | - |
| Sci Rep | No | No | - | Nature | - | - |
| Signa Vitae | No | No | - | Pharmamed Mado | - | - |
| Sleep Breath | No | Yes | Expected | Springer | 11-Jul-22 | 5-Jul-16 |
| Spine | No | No | - | Lippincott W&W | - | - |
| Sports Med | No | Yes | Encouraged | Springer | 10-Aug-20 | - |

| Journal | Evidence synthesis journal | Data policy present | Type of data policy | Publisher | Earliest recorded date of journal's data policy * | Earliest recorded date of publisher's data policy ** |
|---|---|---|---|---|---|---|
| Steroids | No | Yes | Encouraged | Elsevier | 24-Oct-20 | - |
| Surg Obes Relat Dis | No | Yes | Encouraged | Elsevier | 19-Mar-19 | - |
| Surgery | No | Yes | Encouraged | Elsevier | 24-Aug-20 | - |
| Tech Coloproctol | No | No | - | Springer | - | - |
| Thromb Res | No | Yes | Encouraged | Elsevier | 20-May-20 | - |
| Transl Androl Urol | No | No | - | AME Publishing | - | - |
| Updates Surg | No | Yes | Encouraged | Springer | n/a | 5-Jul-16 |
| Urol J | No | Yes | Encouraged | Elsevier OBO Urology and Nephrology RC | 14-Jan-21 | 4-Sep-17 |
| Vaccine | No | Yes | Encouraged | Elsevier | 19-Apr-20 | - |
| Work | No | No | - | IOS Press | - | - |
| World J Surg | No | Yes | Encouraged | Springer | 22-Jun-20 | - |
| World Neurosurg | No | Yes | Encouraged | Elsevier | 18-Apr-21 | 4-Sep-17 |
| Wound Manag Prev | No | No | - | HMP Communications | - | - |
| Wound Repair Regen | No | Yes | Mandatory | Wiley-Blackwell | n/a | 14-Sep-17 |
| Z Rheumatol | No | No | - | Wiley-Blackwell | - | - |

*Note:*

** For journals with a data policy, we used the Wayback Machine tool (https://web.archive.org/) to search for web archives of the Author Instructions page on each journal's website, and recorded the type of data policy based on the version published prior to 1 November 2020.*

***If no archived version of the Author Instructions page prior to 1 November 2020 was found, we recorded the type of data policy based on the earliest version available and verified that the publisher announced the rollout of a universal data policy prior to 1 November 2020 (which supports our assumption that the data policy of the journal had already been in place before 1 November 2020).*

**Appendix S6: Comparison between reviews of health interventions in 2014 vs 2020**

| Item | Median (IQR) or Frequency (%) | |
|---|---|---|
| | 2020* (N=294) | 2014 (N=110) |
| Number of databases searched | 4 (3-5) | 4 (3-5) |
| Number of studies included in review | 13 (8-21) | 13 (7-23) |
| Number of studies in index meta-analysis | 6 (4-10) | 6 (3-11) |
| Country of corresponding author | | |
| China (2014) \| China (2020) | 95/294 (32) | 23/110 (21) |
| UK (2014) \| USA (2020) | 29/294 (10) | 17/110 (15) |
| Canada (2014) \| UK (2020) | 24/294 (8) | 15/110 (14) |
| Other | 146/294 (50) | 55/110 (50) |
| Cochrane reviews | 8/294 (3) | 32/110 (29) |
| Source of funding | | |
| Author reported funding | 115/294 (39) | 60/110 (55) |
| Author reported no funding | 95/294 (32) | 12/110 (11) |
| Not reported | 84/294 (29) | 38/110 (35) |
| Conflict of interest declared | 276/294 (94) | 103/110 (94) |
| Area(s) of intervention | | |
| Pharmacological | 102/294 (35) | 55/110 (50) |
| Non-pharmacological | 183/294 (62) | 43/110 (39) |
| Both | 9/294 (3) | 12/110 (11) |
| ICD-11 category investigated | | |
| Endocrine, nutritional or metabolic diseases (2020)<br>Diseases of the digestive system (2014) | 36/294 (12) | 14/110 (13) |
| Diseases of the digestive system (2020)<br>Infectious or parasitic diseases (2014) | 36/294 (12) | 13/110 (12) |
| Diseases of the musculoskeletal system (2020)<br>Diseases of the circulatory system (2014) | 35/294 (12) | 13/110 (12) |
| Diseases of the circulatory system (2020)<br>Neoplasms (2014) | 30/294 (10) | 11/110 (10) |
| Other | 157/294 (53) | 59/110 (54) |
| Citing a reporting guideline | 242/294 (82) | 32/110 (29) |
| Protocol/registration record cited | | |
| Both protocol and registration record cited | 1/294 (<0.5) | 4/110 (4) |
| Only a protocol cited | 12/294 (4) | 32/110 (29) |
| Only a registration record cited | 110/294 (37) | 2/110 (2) |
| Neither | 171/294 (58) | 72/110 (65) |
| Type(s) of eligible study design stated | 272/294 (93) | 104/110 (95) |
| Randomised studies | 164/272 (60) | 71/110 (65) |
| Non-randomised studies | 20/272 (7) | 7/110 (6) |
| Both | 88/272 (32) | 32/110 (29) |
| Dates of coverage of databases reported | | |
| Both start and end dates | 134/294 (46) | 77/110 (70) |
| Either start or end date only | 152/294 (52) | 29/110 (26) |
| Not reported | 8/294 (3) | 4/110 (4) |
| Boolean search strategy reported for at least 1 database | 211/294 (72) | 60/110 (55) |
| Trials register searched | 63/294 (21) | 38/110 (35) |

| Item | Median (IQR) or Frequency (%) | |
|---|---|---|
| | 2020* (N=294) | 2014 (N=110) |
| Other electronic sources searched | 97/294 (33) | 101/110 (92) |
| Method of study screening reported | 228/294 (78) | 85/110 (77) |
| Method of data collection reported | 227/294 (77) | 84/110 (76) |
| Method of ROB assessment reported | 181/294 (62) | 63/95 (66) |
| Total records retrieved reported | 294/294 (100) | 91/110 (83) |
| Software(s) used for meta-analysis | | |
| R | 33/294 (11) | 7/110 (6) |
| Stata | 73/294 (25) | 16/110 (15) |
| Review Manager | 185/294 (63) | 80/110 (73) |
| Comprehensive Meta-Analysis | 25/294 (9) | 6/110 (5) |
| SAS | 1/294 (<0.5) | 2/110 (2) |
| SPSS | 3/294 (1) | 4/110 (4) |
| Other | 13/294 (4) | 12/110 (11) |
| Not reported | 3/294 (1) | 1/110 (1) |
| Sharing of data or materials used in analyses | 19/294 (6) | 33/110 (30) |
| **Index meta-analysis** | | |
| Measure of effect used | | |
| Risk ratio | 70/294 (24) | 36/110 (33) |
| Odds ratio | 70/294 (24) | 20/110 (18) |
| Hazard ratio | 13/294 (4) | 6/110 (5) |
| Risk difference | 3/294 (1) | 2/110 (2) |
| Mean difference | 76/294 (26) | 25/110 (23) |
| Standardised mean difference | 60/294 (20) | 10/110 (9) |
| Other | 2/294 (1) | 11/110 (10) |
| Method of data preparation reported | 100/294 (34) | 17/110 (15) |
| Meta-analysis model reported (e.g. fixed-effects, random-effects) | 289/294 (98) | 106/110 (96) |
| Summary statistics reported for each study | 211/294 (72) | 79/110 (72) |
| Effect estimate and measure of precision reported for each study | 282/294 (96) | 101/110 (92) |

**The 2020 sample only includes reviews of health interventions.*

### Figure S1: Sensitivity analysis – Frequency of reporting items between systematic reviews published in 2014 and 2020

| Reported item | | 2020 | 2014 | Risk ratio (95% CI) |
|---|---|---|---|---|
| Citation of a reporting guideline | OA | 242/294 (82) | 32/110 (29) | 2.8 (2.1 to 3.8) |
| | SA | 234/286 (82) | 32/78 (41) | 2.0 (1.5 to 2.6) |
| Funding source | OA | 210/294 (71) | 72/110 (65) | 1.1 (0.9 to 1.3) |
| | SA | 204/286 (71) | 42/78 (54) | 1.3 (1.1 to 1.6) |
| Conflict of interest | OA | 276/294 (94) | 103/110 (94) | 1.0 (0.9 to 1.1) |
| | SA | 268/286 (94) | 71/78 (91) | 1.0 (1.0 to 1.1) |
| Protocol / registration record | OA | 123/294 (42) | 38/110 (35) | 1.2 (0.9 to 1.6) |
| | SA | 115/286 (40) | 7/78 (9) | 4.5 (2.2 to 9.2) |
| Eligiblity criteria - Outcomes | OA | 292/294 (99) | 105/110 (95) | 1.0 (1.0 to 1.1) |
| | SA | 284/286 (99) | 73/78 (94) | 1.1 (1.0 to 1.1) |
| Eligibility criteria - Study design | OA | 272/294 (93) | 104/110 (95) | 1.0 (0.9 to 1.0) |
| | SA | 264/286 (92) | 72/78 (92) | 1.0 (0.9 to 1.1) |
| Dates of coverage of databases | OA | 236/294 (80) | 77/110 (70) | 1.1 (1.0 to 1.3) |
| | SA | 229/286 (80) | 48/78 (62) | 1.3 (1.1 to 1.6) |
| Search strategy - Databases | OA | 211/294 (72) | 60/110 (55) | 1.3 (1.1 to 1.6) |
| | SA | 203/286 (71) | 28/78 (36) | 2.0 (1.5 to 2.7) |
| Screening method | OA | 228/294 (78) | 85/110 (77) | 1.0 (0.9 to 1.1) |
| | SA | 220/286 (77) | 53/78 (68) | 1.1 (1.0 to 1.3) |
| Data collection method | OA | 227/294 (77) | 84/110 (76) | 1.0 (0.9 to 1.1) |
| | SA | 219/286 (77) | 53/78 (68) | 1.1 (1.0 to 1.3) |
| ROB assessment method | OA | 181/294 (62) | 63/95 (66) | 0.9 (0.8 to 1.1) |
| | SA | 173/286 (60) | 35/63 (56) | 1.1 (0.9 to 1.4) |
| Total records retrieved | OA | 294/294 (100) | 91/110 (83) | 1.2 (1.1 to 1.3) |
| | SA | 286/286 (100) | 72/78 (92) | 1.1 (1.0 to 1.2) |
| Meta-analysis model (e.g. fixed-effects, random-effects) | OA | 289/294 (98) | 106/110 (96) | 1.0 (1.0 to 1.1) |
| | SA | 281/286 (98) | 74/78 (95) | 1.0 (1.0 to 1.1) |
| Summary statistics per study | OA | 211/294 (72) | 79/110 (72) | 1.0 (0.9 to 1.1) |
| | SA | 203/286 (71) | 50/78 (64) | 1.1 (0.9 to 1.3) |
| Effect estimate and measure of precision per study | OA | 282/294 (96) | 101/110 (92) | 1.0 (1.0 to 1.1) |
| | SA | 274/286 (96) | 69/78 (88) | 1.1 (1.0 to 1.2) |
| Data preparation method | OA | 100/294 (34) | 17/110 (15) | 2.2 (1.4 to 3.5) |
| | SA | 92/286 (32) | 12/78 (15) | 2.1 (1.2 to 3.6) |
| Sharing of data and materials | OA | 19/294 (6) | 33/110 (30) | 0.2 (0.1 to 0.4) |
| | SA | 11/286 (4) | 1/78 (1) | 2.2 (0.4 to 12.0) |

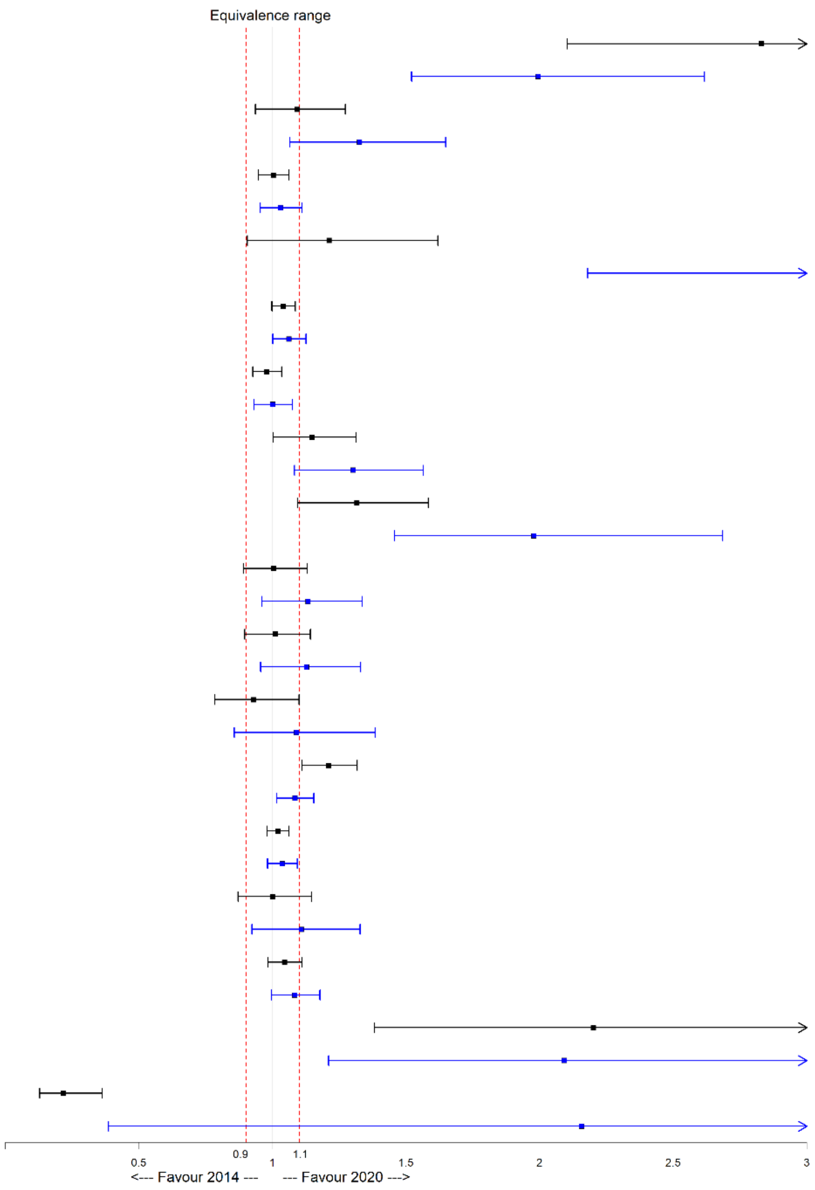

Legend OA Original analysis
SA Sensitivity analysis (excluding Cochrane reviews)

### Figure S2A: Sensitivity analysis – Relationship between citation of reporting guidelines and reported items

| Reported item | | Guideline | No guideline | Risk ratio (95% CI) |
|---|---|---|---|---|
| Funding source | OA | 172/245 (70) | 43/55 (78) | 0.9 (0.8 to 1.1) |
| | SA | 168/239 (70) | 42/53 (79) | 0.9 (0.8 to 1.0) |
| Conflict of interest | OA | 226/245 (92) | 55/55 (100) | 0.9 (0.9 to 1.0) |
| | SA | 221/239 (92) | 53/53 (100) | 0.9 (0.9 to 1.0) |
| Protocol / registration record | OA | 108/245 (44) | 18/55 (33) | 1.3 (0.9 to 2.0) |
| | SA | 107/239 (45) | 17/53 (32) | 1.4 (0.9 to 2.1) |
| Eligibility criteria - Participants | OA | 225/245 (92) | 50/55 (91) | 1.0 (0.9 to 1.1) |
| | SA | 220/239 (92) | 48/53 (91) | 1.0 (0.9 to 1.1) |
| Eligibility criteria - Intervention | OA | 241/245 (98) | 55/55 (100) | 1.0 (1.0 to 1.0) |
| | SA | 236/239 (99) | 53/53 (100) | 1.0 (1.0 to 1.0) |
| Eligibility criteria - Comparator | OA | 219/245 (89) | 48/55 (87) | 1.0 (0.9 to 1.1) |
| | SA | 215/239 (90) | 46/53 (87) | 1.0 (0.9 to 1.2) |
| Eligiblity criteria - Outcomes | OA | 243/245 (99) | 55/55 (100) | 1.0 (1.0 to 1.0) |
| | SA | 237/239 (99) | 53/53 (100) | 1.0 (1.0 to 1.0) |
| Eligibility criteria - Study design | OA | 229/245 (93) | 49/55 (89) | 1.0 (1.0 to 1.2) |
| | SA | 225/239 (94) | 47/53 (89) | 1.1 (1.0 to 1.2) |
| Interface used to search databases | OA | 94/245 (38) | 18/55 (33) | 1.2 (0.8 to 1.8) |
| | SA | 91/239 (38) | 17/53 (32) | 1.2 (0.8 to 1.8) |
| Dates of coverage of databases | OA | 194/245 (79) | 47/55 (85) | 0.9 (0.8 to 1.1) |
| | SA | 190/239 (79) | 46/53 (87) | 0.9 (0.8 to 1.0) |
| Date of last search | OA | 62/245 (25) | 10/55 (18) | 1.4 (0.8 to 2.5) |
| | SA | 57/239 (24) | 9/53 (17) | 1.4 (0.7 to 2.7) |
| Search strategy - Databases | OA | 178/245 (73) | 36/55 (65) | 1.1 (0.9 to 1.4) |
| | SA | 172/239 (72) | 35/53 (66) | 1.1 (0.9 to 1.3) |
| Search strategy - Other sources | OA | 22/115 (19) | 2/25 (8) | 2.3 (0.6 to 9.0) |
| | SA | 22/110 (20) | 2/23 (9) | 2.3 (0.6 to 9.0) |

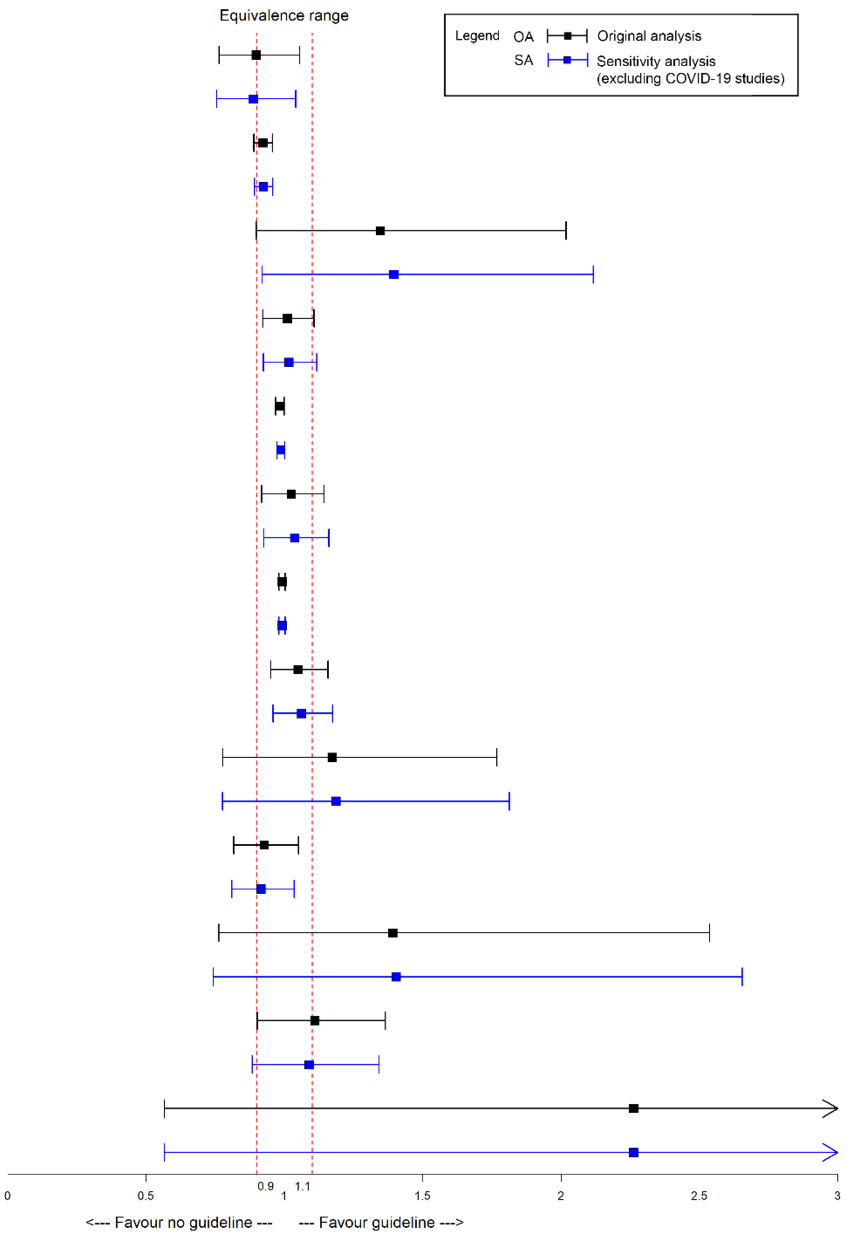

### Figure S2B: Sensitivity analysis – Relationship between citation of reporting guidelines and reported items (ctn'd)

| Reported item | | Guideline | No guideline | Risk ratio (95% CI) |
|---|---|---|---|---|
| Screening method | OA | 195/245 (80) | 38/55 (69) | 1.2 (1.0 to 1.4) |
| | SA | 191/239 (80) | 36/53 (68) | 1.2 (1.0 to 1.4) |
| Data collection method | OA | 182/245 (74) | 47/55 (85) | 0.9 (0.8 to 1.0) |
| | SA | 176/239 (74) | 45/53 (85) | 0.9 (0.8 to 1.0) |
| ROB assessment method | OA | 154/245 (63) | 31/55 (56) | 1.1 (0.9 to 1.4) |
| | SA | 148/239 (62) | 29/53 (55) | 1.1 (0.9 to 1.5) |
| ROB assessment per study | OA | 190/245 (78) | 41/55 (75) | 1.0 (0.9 to 1.2) |
| | SA | 184/239 (77) | 40/53 (75) | 1.0 (0.9 to 1.2) |
| Total records retrieved | OA | 245/245 (100) | 55/55 (100) | 1.0 (1.0 to 1.0) |
| | SA | 239/239 (100) | 53/53 (100) | 1.0 (1.0 to 1.0) |
| Records retrieved per database | OA | 109/245 (44) | 17/55 (31) | 1.4 (0.9 to 2.2) |
| | SA | 106/239 (44) | 17/53 (32) | 1.4 (0.9 to 2.1) |
| Citing at least 1 excluded articles | OA | 56/245 (23) | 9/55 (16) | 1.4 (0.7 to 2.6) |
| | SA | 54/239 (23) | 8/53 (15) | 1.5 (0.8 to 3.0) |
| Data preparation methods | OA | 85/245 (35) | 16/55 (29) | 1.2 (0.8 to 1.9) |
| | SA | 84/239 (35) | 16/53 (30) | 1.2 (0.7 to 1.8) |
| Meta-analysis model (e.g. fixed-effects, random-effects) | OA | 240/245 (98) | 54/55 (98) | 1.0 (1.0 to 1.0) |
| | SA | 234/239 (98) | 52/53 (98) | 1.0 (1.0 to 1.0) |
| Meta-analysis method (e.g. Mantel-Haenszel, inverse variance) | OA | 179/245 (73) | 39/55 (71) | 1.0 (0.9 to 1.2) |
| | SA | 174/239 (73) | 38/53 (72) | 1.0 (0.8 to 1.2) |
| Heterogeneity variance estimator (e.g. DerSimonian Laird) | OA | 45/197 (23) | 5/38 (13) | 1.8 (0.7 to 4.7) |
| | SA | 45/192 (23) | 5/36 (14) | 1.8 (0.7 to 4.7) |
| Summary statistics per study | OA | 174/245 (71) | 41/55 (75) | 1.0 (0.8 to 1.1) |
| | SA | 168/239 (70) | 39/53 (74) | 1.0 (0.8 to 1.1) |
| Effect estimate and measure of precision per study | OA | 234/245 (96) | 54/55 (98) | 1.0 (0.9 to 1.0) |
| | SA | 228/239 (95) | 52/53 (98) | 1.0 (0.9 to 1.0) |
| Sharing of data and materials | OA | 18/245 (7) | 2/55 (4) | 1.7 (0.4 to 6.7) |
| | SA | 18/239 (8) | 2/53 (4) | 1.7 (0.4 to 6.7) |

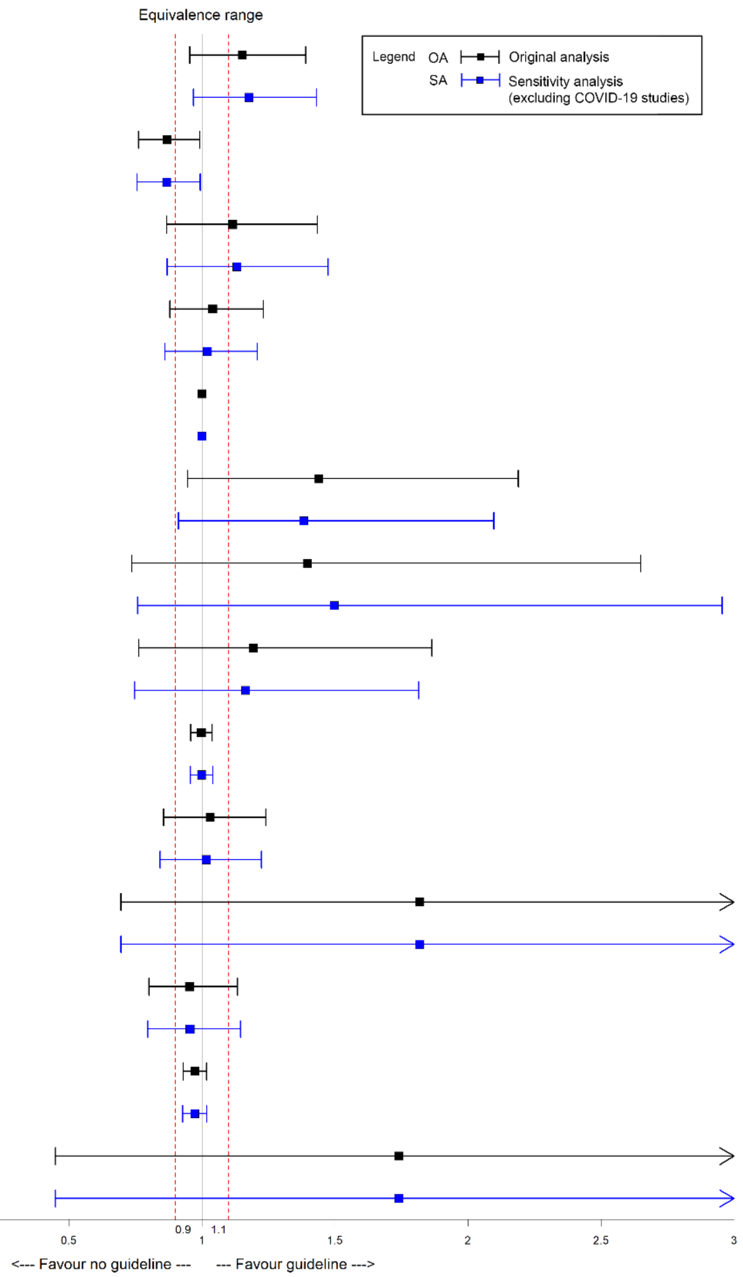

**Figure S3: Relationship between journal's presence of a data sharing policy and reported items**

| Reported item | Presence of data sharing policy | No. of studies reporting (%) | Risk ratio (95% CI) |
|---|---|---|---|
| All search strategies | Y | 59/199 (30) | 1.4 (0.9 to 2.1) |
| | N | 22/101 (22) | |
| Sharing of other data/material | Y | 18/199 (9) | 4.1 (1.1 to 15.5) |
| | N | 2/101 (2) | |
| Unprocessed extracted data | Y | 8/199 (4) | 3.0 (0.5 to 17.2) |
| | N | 1/101 (1) | |
| Data conversions performed | Y | 1/199 (1) | 1.5 (0.1 to 38) |
| | N | 0/101 (0) | |
| Data used in analysis | Y | 12/199 (6) | 13.5 (0.8 to 230.9) |
| | N | 0/101 (0) | |
| Analytic code | Y | 1/199 (1) | 0.5 (0.1 to 4.9) |
| | N | 1/101 (1) | |
| Citations of included & excluded studies | Y | 2/199 (1) | 2.6 (0.1 to 54) |
| | N | 0/101 (0) | |
| Metadata of shared files | Y | 1/199 (1) | 1.5 (0.1 to 38) |
| | N | 0/101 (0) | |

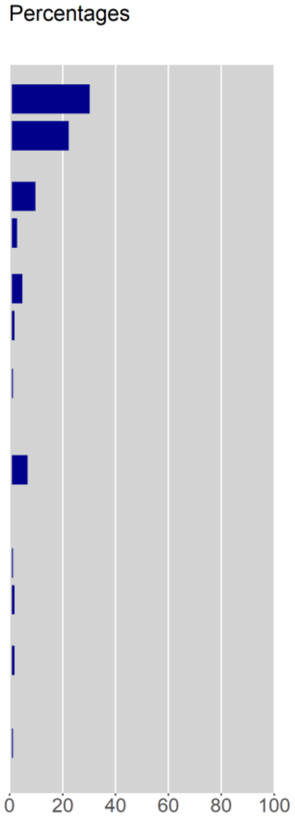

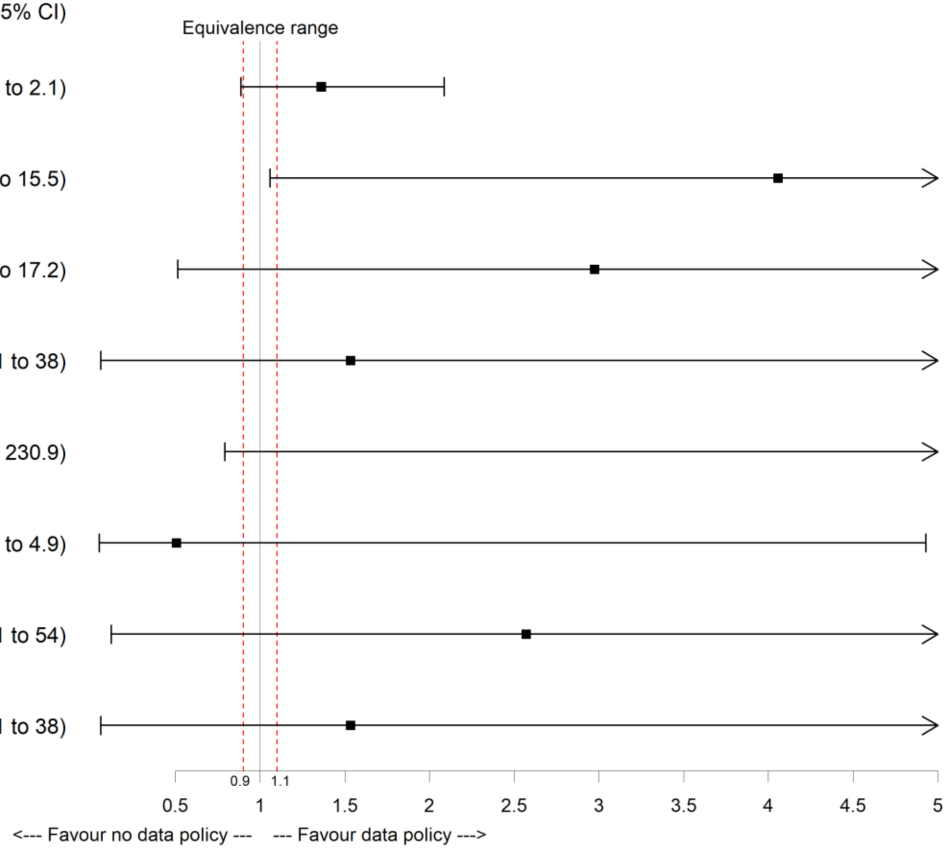
